# Controlling for confounds in UK Biobank brain imaging data with small subsets of subjects

**DOI:** 10.64898/2026.03.02.26347455

**Authors:** Lav Radosavljević, Stephen M. Smith, Thomas E. Nichols

## Abstract

The UK Biobank (UKB) Brain Imaging cohort contains data from almost 100,000 subjects and has yielded invaluable understanding of the links between the brain and health outcomes and lifestyles. Much of the understanding of these links has come from exploring the association between Imaging Derived Phenotypes (IDPs) and other variables that are unrelated to brain imaging, so called non-Imaging Derived Phenotypes (nIDPs). When performing analysis of this kind, it is very important to control for well known confounding factors such as age, sex and socio-economic status, as well as confounds which are related to the imaging protocol itself. In previous work, we created a pipeline for constructing imaging confounds for use in statistical inference via a standard multivariate linear regression approach (Alfaro-Almagro et. al. 2021). However, this approach is problematic when the number of confounds exceeds the number of subjects, and is severely underpowered when the number of number of subjects is not much larger than the number of confounds. In this work, we perform a simulation study to evaluate 13 modelling approaches to account for confounds when their number is similar to or exceeds the number of subjects. Based on the simulation results, we recommend a ridge regression based permutation test for low sample sizes (*n* ≤ 50), a version of de-sparsified LASSO for intermediate sample sizes (50 < *n* ≤ 500), and multivariate linear regression aided by Principal Component Analysis (PCA) for larger sample sizes (*n >* 500). We also demonstrate the use of our recommended methodology on a real data example of finding associations between Alzheimer’s Disease (AD) and IDPs.

## 1 Background

The UK Biobank brain imaging cohort has recently scanned its 100,000-th subject and researchers now have access to data well over 85, 00 subjects. This resource has, among other things, yielded invaluable understanding of the links between imaging and non-imaging variables (Douaud et al., 2022, Dutt et al., 2022, Elliott et al., 2017, Topiwala et al., 2021, 2023). When establishing such links, it is crucial to control for confounding factors to avoid the detection of spurious associations. In Alfaro-Almagro et al. (2021), a set of imaging confounds are identified that are predictive of Imaging Derived Phenotypes (IDPs) and/or non-Imaging Derived Phenotypes (nIDPs). The following regression equation describes the assumed relationship between some univariate exposure vector **x**, univariate outcome vector **y** and confound matrix **C**:

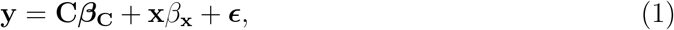

where ***ϵ*** is assumed to be homoscedastic, uncorrelated white noise with Var(*ϵ*_*i*_) = *σ*^2^. In neuroimaging research, **x** is usually an nIDP and **y** an IDP or vice versa. We are interested in testing null-hypotheses on the form *H*_0_: *β*_**x**_ = 0 against *H*_1_: *β*_**x**_ ≠ 0. The most common approach to performing this test is by way of a standard t-test on the coefficient 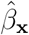 obtained by Ordinary Least Squares (OLS) regression. This simple approach is appropriate when the sample size *n* is equal to the full number of subjects in the UKB Brain imaging cohort, but when working on a subset of all subjects, which is common in neuroimaging research, this approach can be problematic in several ways. Firstly, this approach is not at all possible when *n* < *p*, where *p* is the number of confounds. Secondly, if *n* is is not much larger than *p*, i.e., if the design matrix of the OLS regression is close to square-shaped, the standard approach can be severely underpowered. This is because the variance of 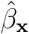 is

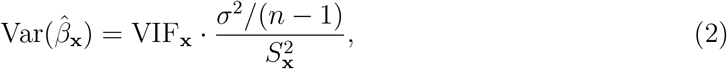

where 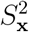 is the sample variance of **x**, and VIF_**x**_ is the Variance Inflation Factor (VIF) of **x**. In other words, the variance of 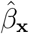 is proportional to VIF_**x**_. The VIF has the following value:

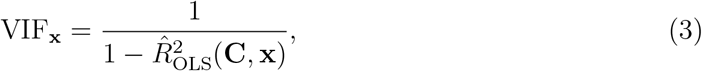

where 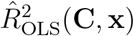 is the proportion of variance explained when regressing **x** on **C**. Since the proportion of variance explained is severely inflated when *n* is not much larger than *p*, this will lead to high standard errors for 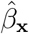, which in turn leads to low power. Several approaches have been formulated to tackle these problems. These methods typically assume some level of sparsity in the coefficients ***β***_**C**_. In this work, we describe a number of these methods and evaluate them using simulated exposure and outcome variables, and real confound data for different sample sizes, levels of sparsity of *β*_*C*_ and levels of partial correlation between outcome and exposure. The goal is to find the method with the highest statistical power, while having a controlled False Positive Rate (FPR). We also show an illustrative example of the use of the most suitable method from the simulation study on real data, by performing mass univariate testing of the association between IDPs and Alzheimer’s Disease (AD).

### 1.1 UKB Imaging Confounds

UK Biobank Brain Imaging confounds are constructed to control for nuisance effects in the IDPs caused by the participants physical and demographic attributes, the technology and protocol used for imaging and miscellaneous drift effects over time. These confounds include, among other things, sex, age, site, head-motion and scanner-protocol. Non-linear transformations of confounds and interaction terms are also considered to ensure that as much nuisance variance as possible is captured. These confounds are also correlated with nIDPs (Alfaro-Almagro et al., 2021), which means that they are more than just nuisance variables which cause regression dilution; if not controlled for, they cause detection of spurious associations between IDPs and nIDPs.

## 2 Methods

In this sub-section, we describe a range of methods that are used for high-dimensional inference that we consider as candidates for use on UKB Brain Imaging Data.

### 2.0.1 Principal Component Regression

Using Principal Component Analysis (PCA) to reduce the dimensionality of confound data is an approach commonly used in Genome Wide Association Studies (GWAS), where principal components of the kinship matrix are included in multivariate regression models to control for confounding effects caused by relatedness of the participants (Thomson and McWhirter, 2017). We include two versions of this approach, `PCA-OLS1` and `PCA-OLS2`, in our simulation study. `PCA-OLS1` uses the following procedure for high-dimensional inference:

1. Perform PCA on confound matrix **C**.
2. Include Principal Components (PCs) by the size of corresponding singular values in an OLS model with outcome **y** as dependent variable and exposure **x** as one of the independent variables. Stop including PCs once the model has only 20 error degrees of freedom remaining, or there are no more PCs remaining to include.
3. Use the standard OLS estimator on this variable set to test the hypothesis *H*_0_: *β*_**x**_ = 0

This is our baseline high-dimensional inference approach, as it is as close to the default OLS approach as possible. Indeed, for settings where *n* − *p* − 2 ≥ 20, this approach is exactly equivalent to OLS. `PCA-OLS2` is a less conservative approach and uses the following procedure:

1. Perform PCA on confound matrix **C**.
2. Include Principal Components (PCs) by the size of corresponding singular values in an OLS model with outcome **y** as dependent variable and exposure **x** as one of the independent variables. Stop including PCs once the PCs explain 95% of the variance in **C**, or when the model reaches 20 remaining error degrees of freedom.
3. Use the standard OLS estimator on this variable set to test the hypothesis *H*_0_: *β*_**x**_ = 0

`PCA-OLS2` is less conservative than `PCA-OLS1`, as it includes a lower number of PCs in the multivariate regression. Both approaches rely to different extents on the *manifold hypothesis*, i.e., that our high-dimensional confound data has a latent low dimensional structure that we can discover using PCA (Whiteley et al., 2022).

### 2.0.2 Uncorrected Ridge Regression p-values

Coefficient estimates of the Ridge Regression (RR) estimator are biased and can therefore not be directly used for exact parametric inference. However, under certain conditions, the bias in the estimator becomes negligible compared to its variance under the null hypothesis, giving us coefficient t-tests that are close to unbiased (Halawa and El Bassiouni, 2000). We use approximate t-test detailed in Cule et al. (2011). Assuming we have a linear model

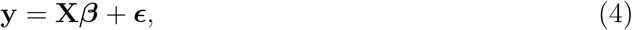

where ***ϵ*** is white noise with variance *σ*^2^, the null hypothesis *H*_0_: *β*_*j*_ = 0 is tested using the following approximate t-statistic:

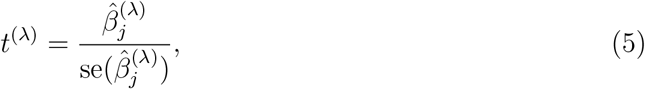

where 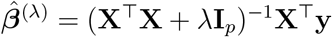 is the RR estimate of ***β*** using penalty *λ*, and 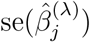 is the square root of the *j*th diagonal element of the matrix

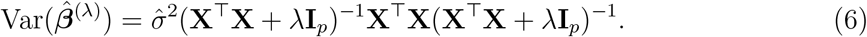

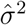 is estimated in the following way:

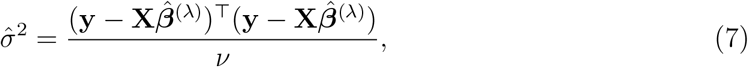

where the error degrees of freedom *ν* is set to be *ν* = *n*− trace(2**H** − **HH**^⊤^) for “hat matrix” **H** = **X**(**X**^⊤^**X** + *λ***I**_*p*_)^−1^**X**^⊤^. *t*^(*λ*)^ is assumed to approximately follow Student’s t-distribution with degrees of freedom *n* − trace(**H**).

The most appropriate method of choosing penalty parameter *λ* is still a hotly debated subject in the scientific community (Ding et al., 2018). We allow for two selection methods, one proposed in Cule and De Iorio (2013) and a standard approach by selecting *λ* through 5-fold Cross-Validation. The two methods are referenced as `Ridge.Cule-DeIorio` and `Ridge.CV` respectively.

### 2.0.3 Ridge Regression Permutation Testing

Hemerik et al. (2021) outline three different RR-based permutation tests for confound adjusted association. The first two tests are based on the Freedman-Lane permutation testing procedure (Freedman and Lane, 1983) and the third test is based on a permutation test by Kennedy where both outcome and exposure are residualised with respect to confounds (Winkler et al., 2014). Each method is described in detail in the original article, but we will only go through the first Freedman-Lane based method `Ridge.PT.FLH1`. Assuming the model described in equation 1, we select penalty term *λ*_**x**_ for regression of **x** on **C** and *λ*_**y**_ for regression of **y** on **C** using cross-validation. The absolute sample correlation between the residuals of both regressions is estimated:

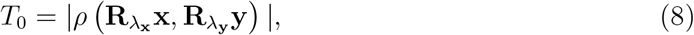

where 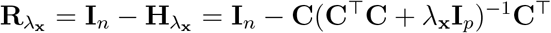 and 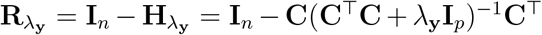. For *b* subsequent permutations **P**_1_, …, **P**_*b*_, the following statistic is calculated

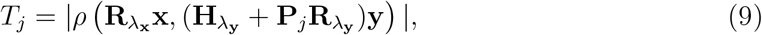

for *j* = 1, …, *b*. The estimated p-value is

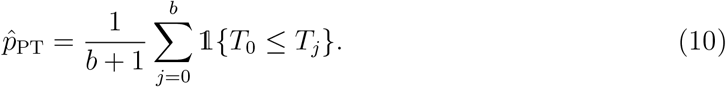

The two methods based on the Freedman-Lane procedure are referred to as `Ridge.PT.FLH1` and `Ridge.PT.FLH2`, and the one based on double residualisation is referred to as `Ridge.PT.DR`.

### 2.0.4 Naive Feature Selection

Feature selection is a common pre-processing step to reduce the dimensionality of inference tasks. The post-selection estimator using the Least Absolute Selection and Shrinkage Operator (LASSO) has been proposed as a viable option for high dimensional inference tasks (Zhao et al., 2021). It works in the following way:

1. Fit a LASSO regression model predicting **y** using **C**.
2. Create a reduced set of confounds 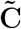, containing only variables that have non-zero coefficient estimates in the LASSO regression.
3. Fit an OLS model regressing **y** on 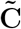 and **x**. If the model has insufficient degrees of freedom, i.e., lower than 20, we use PCA on 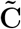 as in sub-section 2.0.1.

Since LASSO is sometimes not appropriate for strongly correlated data, and there are confounds in our data set that are highly correlated, we also use ElasticNet to perform selection instead of LASSO. We call the two methods `Naive.LASSO` and `Naive.ElasticNet`. All tuning parameters are selected via cross-validation.

### 2.0.5 Double Selection

Belloni et al. (2014) propose a post-selection estimator where variables that are predictive of **x** and/or **y** are selected, likewise using LASSO. The selection process works in the following way:

1. Fit a LASSO regression model predicting **x** using **C**.
2. Fit a LASSO regression model predicting **y** using **C**.
3. Create a reduced set of confounds 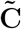, containing only variables that have non-zero coefficient estimates in either LASSO regression.
4. Fit an OLS model regressing **y** on 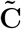 and **x**. If the model has insufficient degrees of freedom, i.e., lower than 20, we use PCA on 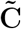 as in sub-section 2.0.1.

It is very similar to the naive selection estimator, but with two key advantages. Firstly, it increases power by more strongly orthogonalising **x** with respect to confounds as we include more confounds that are highly predictive of **x**. Secondly, FPR control is improved by including variables that are strongly predictive of **x** and weakly predictive of **y**. Failure to include such variables in the single selection estimator (see sub-section 2.0.4) leads to inflation in the FPR.

We refer to this method as LASSO Double Selection (`LADS`). As with the single selection estimator, we might expect to see some benefit in using ElasticNet over LASSO, and therefore also include an estimator which we refer to as ElasticNet Double Selection (`ENDS`).

### 2.0.6 De-Sparsified LASSO

De-Sparsified or de-biased LASSO is a process by which bias in the LASSO estimator is corrected so that inference can be performed on the parameter estimates (Chevalier, 2020). Assuming a standard linear model as in equation 4, Javanmard and Montanari (2014) propose the following estimator:

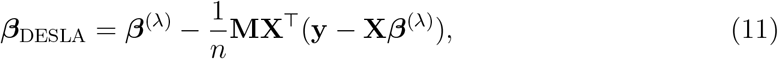

where ***β***^(*λ*)^ is a LASSO coefficient estimate and **M** is an estimate of the inverse of the covariance matrix of **X**. It is easy to see intuitively why this correction generally reduces the bias in the LASSO estimate ***β***^(*λ*)^. If 𝔼[***β***^(*λ*)^] = ***β*** + ***γ***, i.e., ***β***^(*λ*)^, has bias ***γ***, then 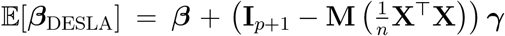. Since **M** is an approximation of the precision matrix of **X**, it follows that 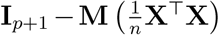 will be close to the null matrix, thus reducing the bias. The estimator has approximately the following covariance matrix:

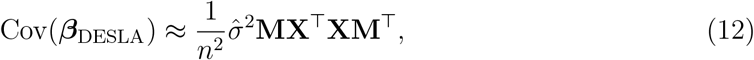

where 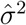 is an estimate of the residual variance.

The choice of method for finding *λ* and **M** are crucial details in constructing this estimator. Javanmard and Montanari propose a convex program for determining **M** and a heuristic for determining *λ* which are the default settings in their `R` function `SSLasso`. A much faster and more conservative version of this estimator has been proposed in Boot and Nibbering (2017) that uses the Moore-Penrose pseudo-inverse of 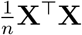 as **M**. For this version of the de-sparsified LASSO estimator, we use cross-validation to select *λ*. The two methods are referred to as `DESLA-JM` and `DESLA-MP`. De-sparsified LASSO is generally not used when *n* > *p*, so we limit the use of these methods to scenarios where *n* < *p*, in the simulation study.

#### 2.1 Simulation Method

We are interested in the FPR control and power of all estimators under different levels of sparsity, sample size and strength of confounding. We design a simulation framework that uses real, standardised UKB imaging confound data **C** with simulated outcomes **y** and exposures **x**. Specifically, we assume the following system of regression equations for describing **x** and **y**:

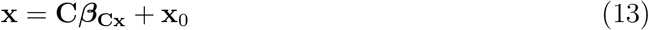

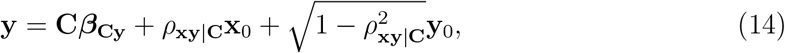

where **x**_0_ ∼ 𝒩 (**0, I**_*n*_) and **y**_0_ ∼ 𝒩 (**0, I**_*n*_) are independent from each other and all other variables. Here, **C*β***_**Cx**_ and **C*β***_**Cy**_ are the confounding signals that go into **x** and **y** respectively, **x**_0_ is the variance component in **x** that is independent from confounds, and **y**_0_ is the variance component in **y** that is independent from both confounds and **x**. Since **x**_0_ and **y**_0_ are simulated to have unit variance, *ρ*_**xy**|**C**_ is the partial correlation between **x** and **y** after adjusting for confounds, which is precisely the association that we wish to test.

***β***_**Cx**_ and ***β***_**Cy**_ are also simulated to have varying degrees of approximate sparsity and correlation between each other. The reason we are also interested in sparsity is that the imaging confounds are unlikely to all be equally relevant to each variable of interest, and this can be leveraged to increase inference power. We induce correlation between ***β***_**Cx**_ and ***β***_**Cy**_, because some variables, such as age, are likely to be strongly associated with both outcome and exposure, which makes it unrealistic to simulate them independently. Specifically, for each confound *j*, we simulate

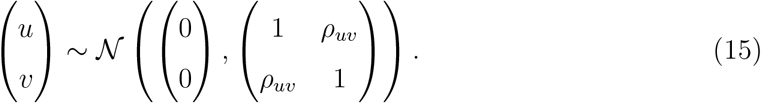

We then set *β*_**Cx**,*j*_ = sign(*u*_*j*_)|*u*_*j*_|^*δ*^ and *β*_**Cy**,*j*_ = sign(*v*_*j*_)|*v*_*j*_|^*δ*^, where *δ* is a positive integer. Depending on the size of *δ*, this introduces some controlled level of approximate sparsity in ***β***_**Cx**_ and ***β***_**Cy**_, where a higher *δ* means higher sparsity. The correlation *ρ*_*uv*_ is mathematically calibrated so that corr(*β*_**Cx**,*j*_, *β*_**Cy**,*j*_) = *ρ*_*β*_, for some pre-chosen *ρ*_*β*_. A higher value of *ρ*_*β*_ is more likely to cause false positives, since it means that more confounds are strongly predictive of both outcome and exposure. ***β***_**Cx**_ and ***β***_**Cy**_ are also normalised so that the confounds explain 25% of variance in both **x** and **y**.

The number of imaging confounds that we use is *p* = 754 and the sample size *n* is varied to be *n* = 50, 100, 200, 500, 1000, 2000. The sparsity parameter *δ* is varied to be *δ* = 1, 2, 3, 5, 7, 10, and *ρ*_*β*_ is varied to be *ρ*_*β*_ = 0, 0.25, 0.5, representing weak, intermediate and strong confounding. Finally if *n* ≤ 1000, we vary *ρ*_**xy**|**C**_ to be *ρ*_**xy**|**C**_ = 0, 0.05, 0.1, 0.2, 0.4, and if *n* = 2000, we let *ρ*_**xy**|**C**_ = 0, 0.01, 0.02, 0.05, 0.1. This is because smaller effect sizes are more relevant to the setting of *n* = 2000. *ρ*_**xy**|**C**_ = 0 means that there is no association between **x** and **y** after adjusting for confounds and is the setting that is used to evaluate the FPR control of our methods. Each setting is evaluated over 1000 random simulations to calculate power and FPR.

Our simulation framework differs from previous work on the topic, as it produces approximate sparsity in confounding effects ***β***_**C**_, rather than exact sparsity (Hemerik et al., 2021, Zhao et al., 2021). We see this as a more realistic setting than exact sparsity, as confounding effect sizes are unlikely to be truly 0 in reality, but may vary heavily in magnitude. Most previous simulation studies also treat the confounding effects as block homogeneous, i.e., that large blocks of confounds have the exact same effect. This differs significantly from real data, where different confounds have very different effect sizes.

#### 2.2 Real Data Demonstration

To demonstrate the use of high-dimensional inference methodology on real data, we perform mass univariate testing between 3939 IDPs and one nIDP, adjusting for all imaging confounds. Here, the IDPs are modelled as outcome and the nIDP as exposure. The nIDP that we choose is AD and the data consists of 64 AD subjects and 128 age- and sex-matched controls, giving us a sample size of *n* = 192 in total. The mass univariate analyses are also performed using the baseline method, `PCA-OLS1`, for comparison.

## 3 Results

### 3.1 Simulation Results

The simulation results are too extensive to present in the main text of this paper, considering that we evaluate 6 · 6 · 3 · 5 = 540 simulation settings. We therefore focus on a relatively small subset of simulation settings. Being conservative, we fix *ρ*_*β*_ = 0.5, which is the setting most prone to cause false positives for all methods. We vary the sample size to be *n* = 50, 100, 200, 500, 1000, 2000, covering all sample sizes of interest, and we vary the sparsity parameter *δ* to be *δ* = 1, 3, 7, representing a dense, medium and sparse setting. The results for the same simulation settings but with *ρ*_*β*_ = 0, 0.25 are in the supplementary material (see 5).

The results of the simulation study are shown in Figures 1 through 6, each showing the results for a given number of subjects *n*, with varying sparsity *δ* = 1, 3, 7. For *n* = 50 subjects, we see that `PCA-OLS1`, `PCA-OLS2` and `Ridge.PT.FLH1` seem to have reasonably controlled FPRs, with `Ridge.PT.FLH1` having a somewhat conservative FPR, and also the highest power. For *n* = 100 subjects, we see that `PCA-OLS1`, `PCA-OLS2` and `DESLA-MP` are the only methods that have controlled FPR. Of these three, `DESLA-MP` has the highest power. For *n* = 200, the same three methods are the only ones with controlled FPR, but `PCA-OLS2` now has the highest power, followed by `DESLA-MP`. Also, we see that `DESLA-MP` has a conservative FPR compared to `PCA-OLS1` and `PCA-OLS2`. Additionally, we see that the methods based on double selection, `LADS` and `ENDS`, are close to having a controlled FPR for the most sparse setting *δ* = 7. The results follow a very similar pattern for *n* = 500, where the same three methods have a controlled FPR and `PCA-OLS2` has by far the highest power, followed by `DESLA-MP`. Here, we also clearly see a conservative FPR for `DESLA-MP` compared to `PCA-OLS1` and `PCA-OLS2`. Also for the highest sparsity, we now have a fully controlled FPR for the two methods based on double selection. For *n* = 1000, `OLS-PCA1` is the only method with a fully controlled FPR in all scenarios (for *n* = 1000, it is equivalent to regular OLS), with `OLS-PCA2` and `ENDS` showing borderline controlled FPRs and high power. `LADS` also seems to have a controlled FPR, except for in the most dense scenario, *δ* = 1. For *n* = 2000, we now see that only `OLS-PCA1` and `ENDS` have controlled FPRs, but with fairly similar power.

**Figure 1.**
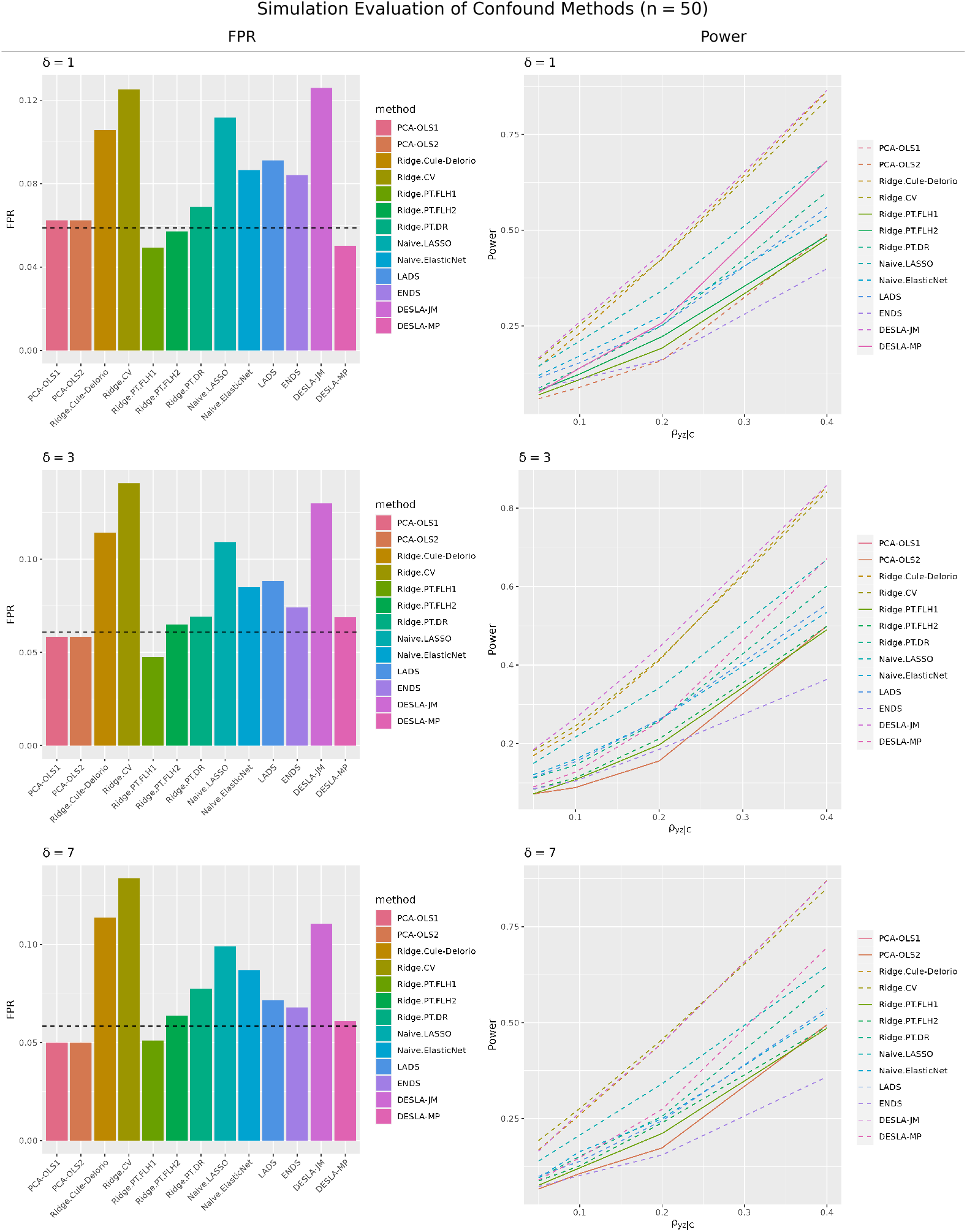
Simulation results for *n* = 50. The bar plots to the left show the FPR and lineplots the statistical power of different methods. The dashed line in the bar plots is the 95% upper confidence limit for a FPR of 0.05. Methods above this limit have dashed lines in the power plot. Here, we see that methods `PCA-OLS1`, `PCA-OLS2` and `Ridge.PT.FLH1` have reasonably controlled FPRs, with `DESLA-MP` having the highest power.

**Figure 2.**
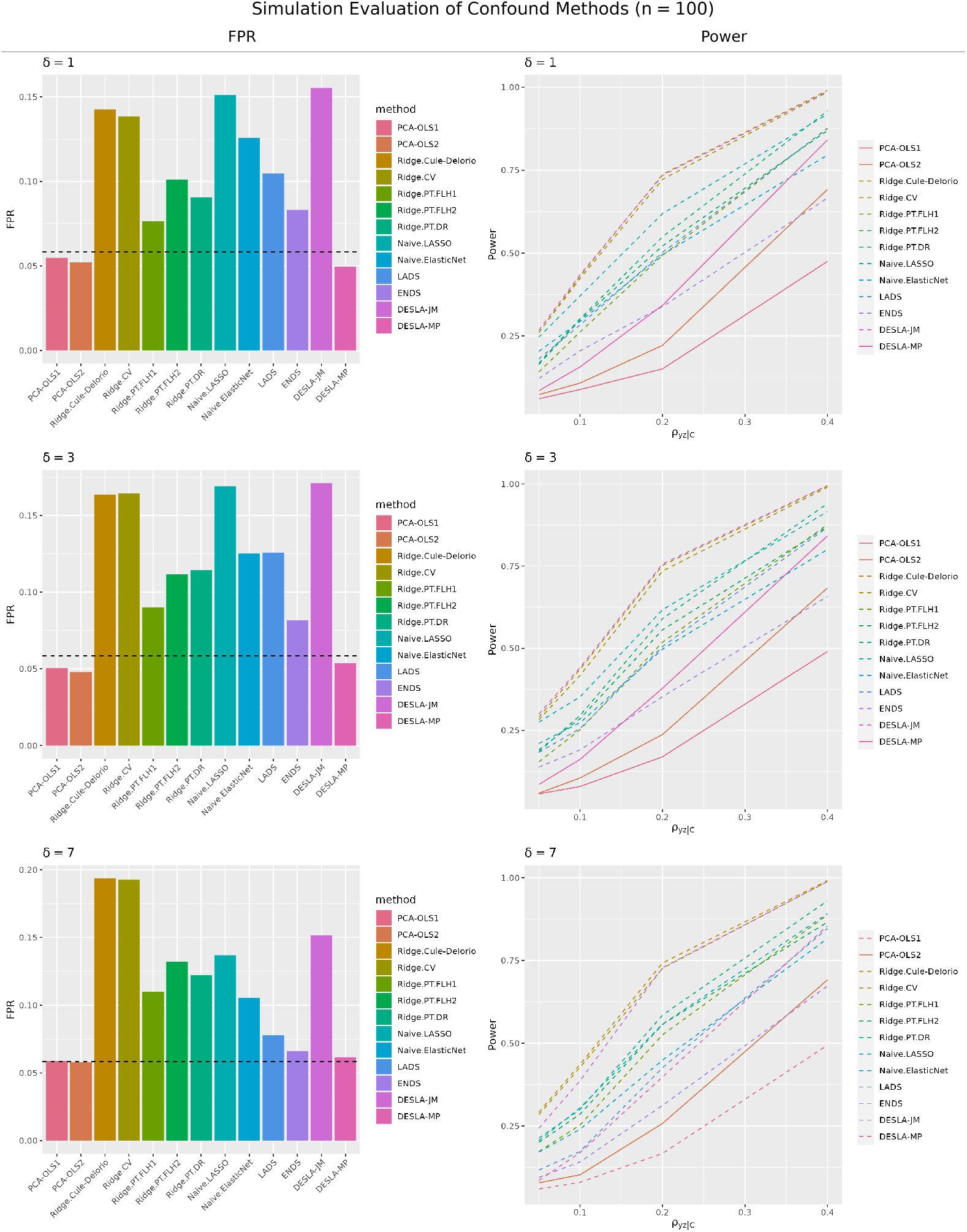
Simulation results for *n* = 100. The bar plots to the left show the FPR and lineplots the statistical power of different methods. The dashed line in the bar plots is the 95% upper confidence limit for a FPR of 0.05. Methods above this limit have dashed lines in the power plot. Here, we see that methods `PCA-OLS1`, `PCA-OLS2` and `DESLA-MP` have reasonably controlled FPRs, with `DESLA-MP` having the highest power.

**Figure 3.**
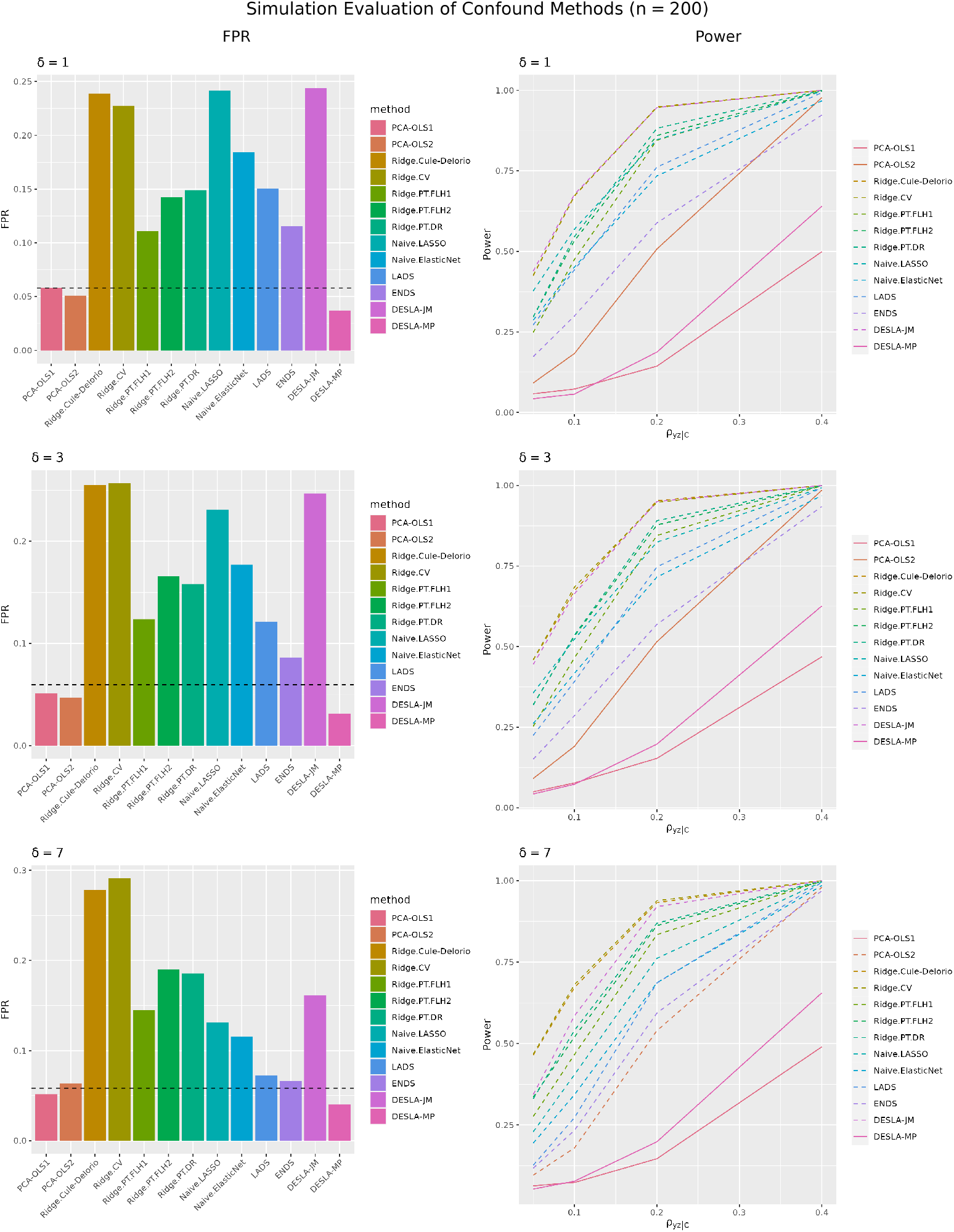
Simulation results for *n* = 200. The bar plots to the left show the FPR and lineplots the statistical power of different methods. The dashed line in the bar plots is the 95% upper confidence limit for a FPR of 0.05. Methods above this limit have dashed lines in the power plot. Here, we see that methods `PCA-OLS1, PCA-OLS2` and `DESLA-MP` have reasonably controlled FPRs, with `PCA-OLS2` having the highest power. `DESLA-MP` is shown to have a conservative FPR.

**Figure 4.**
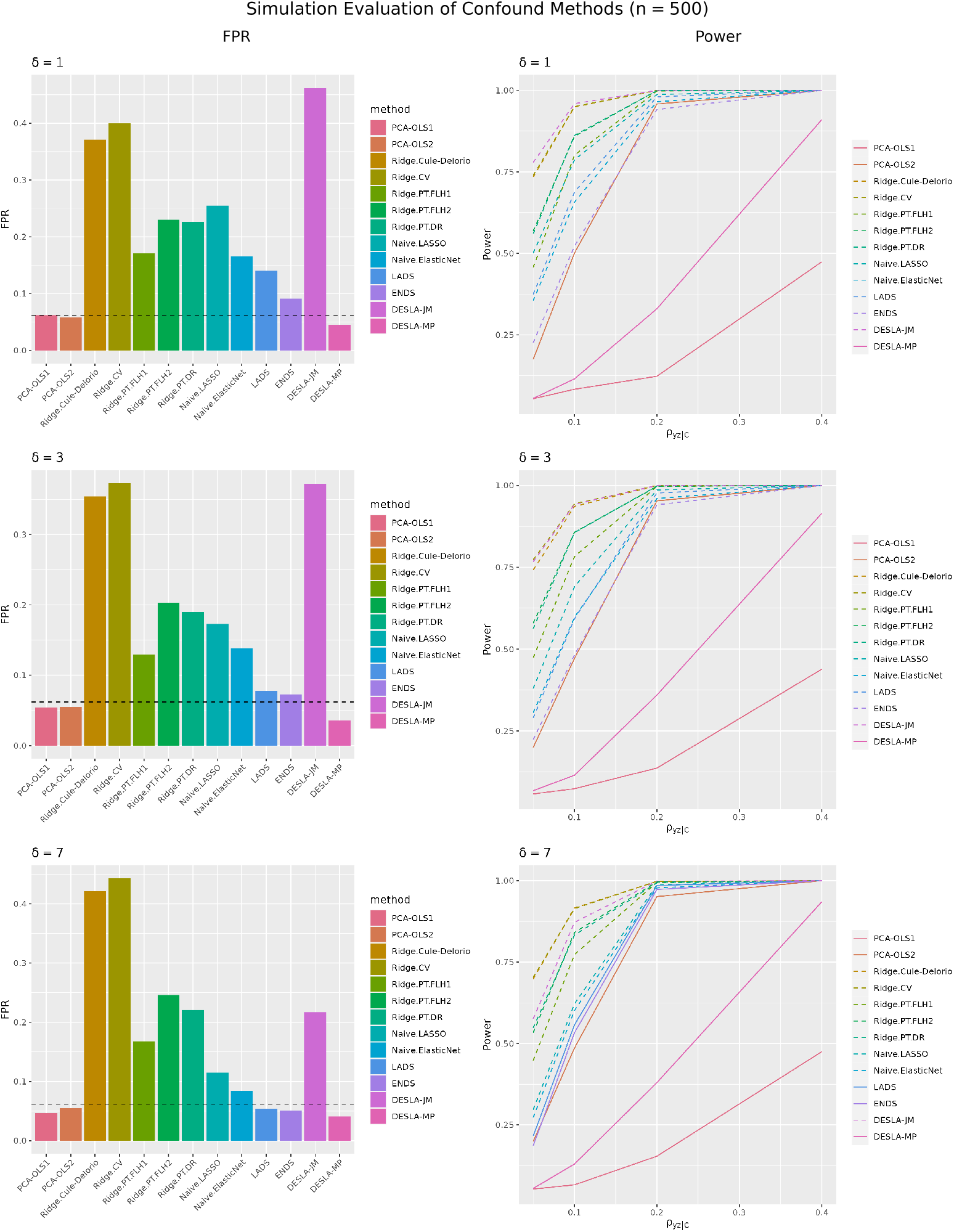
Simulation results for *n* = 500. The bar plots to the left show the FPR and lineplots the statistical power of different methods. The dashed line in the bar plots is the 95% upper confidence limit for a FPR of 0.05. Methods above this limit have dashed lines in the power plot. Here, we see that methods `PCA-OLS1, PCA-OLS2` and `DESLA-MP` have reasonably controlled FPRs, with `PCA-OLS2` having the highest power. `DESLA-MP` is shown to have a conservative FPR.

**Figure 5.**
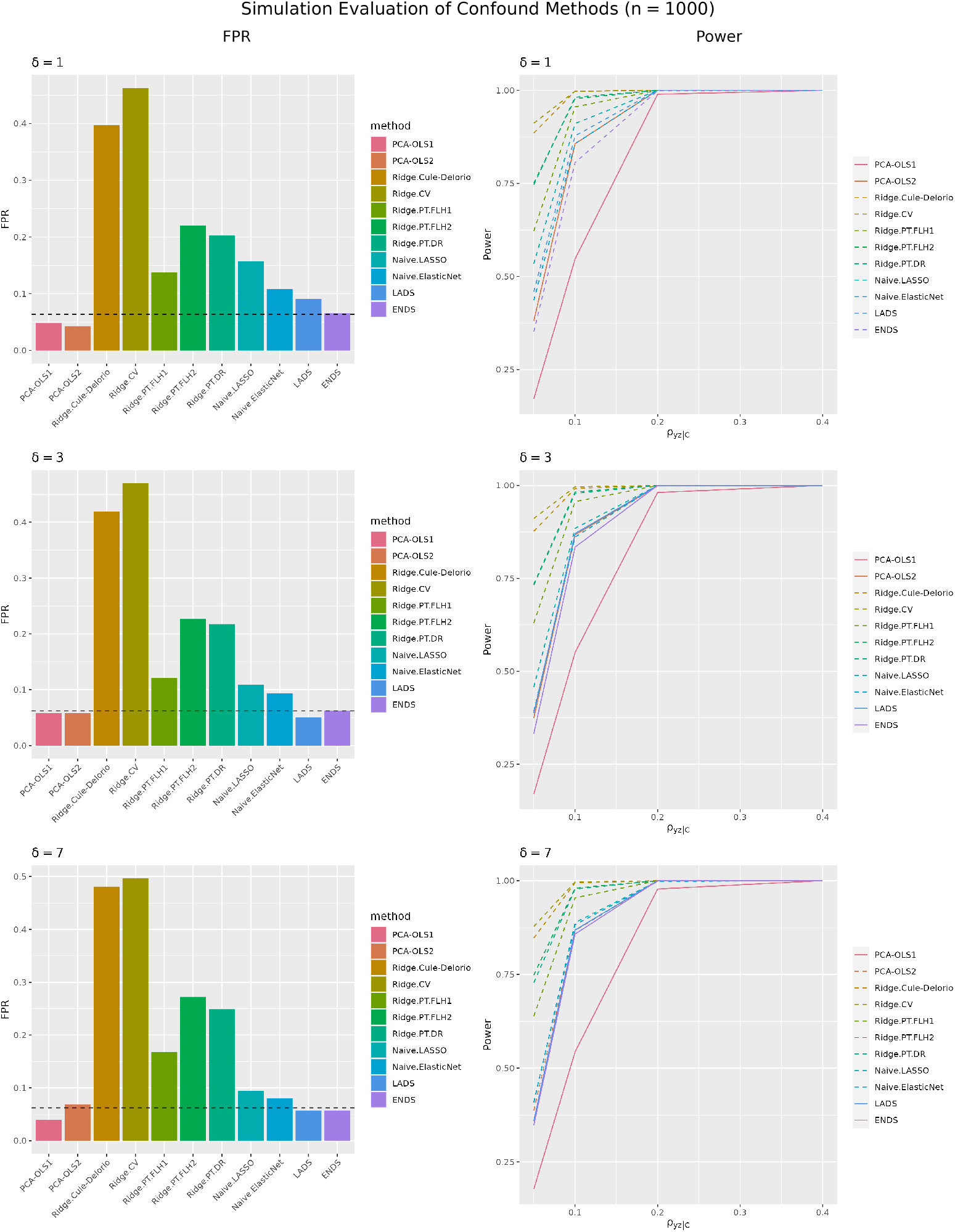
Simulation results for *n* = 1000. The bar plots to the left show the FPR and lineplots the statistical power of different methods. The dashed line in the bar plots is the 95% upper confidence limit for a FPR of 0.05. Methods above this limit have dashed lines in the power plot. Here, we see that only `PCA-OLS1` has a reasonably controlled FPRs, with `PCA-OLS2` and `ENDS` having borderline controlled FPRs and higher power than `PCA-OLS1`.

**Figure 6.**
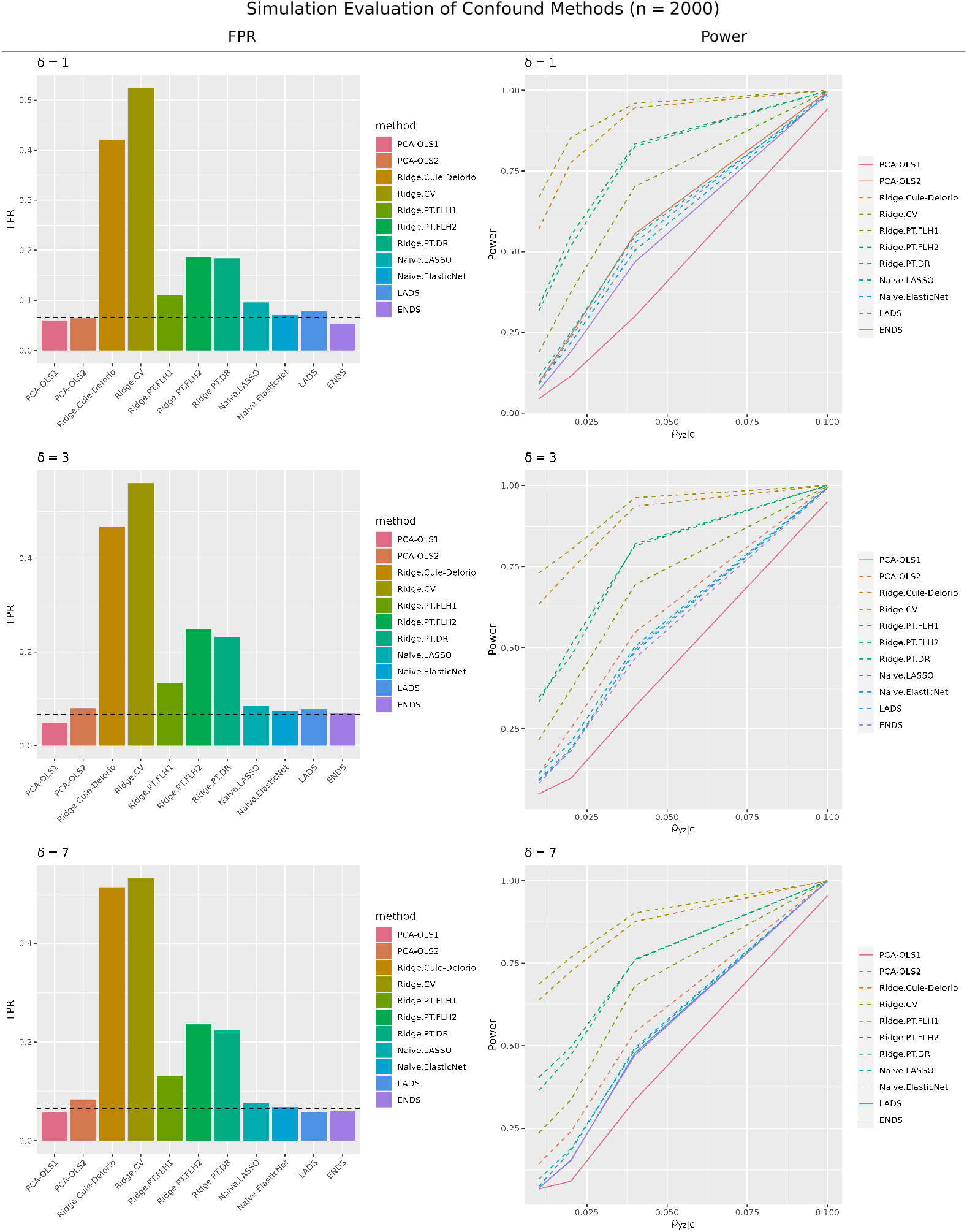
Simulation results for *n* = 2000. The bar plots to the left show the FPR and lineplots the statistical power of different methods. The dashed line in the bar plots is the 95% upper confidence limit for a FPR of 0.05. Methods above this limit have dashed lines in the power plot. Here, we see that only `PCA-OLS1` and `ENDS` have controlled FPRs, but with quite similar statistical power.

From these results, it is clear that there are suitable methods for high-dimensional inference in UK Biobank neuroimaging research at every level of sample size. Our real data task of testing the association between IDPs and AD, has sample size *n* = 192, which is closest to *n* = 200. In this setting, we see that `PCA-OLS1` and `DESLA-MP` both have higher power than `PCA-OLS1`, but that `PCA-OLS1` has a slightly elevated empirical FPR for *δ* = 7, while `DESLA-MP` has a conservative FPR. We therefore choose to use `DESLA-MP` for the exemplar study, comparing the performance of this method with the baseline approach, `PCA-OLS1`.

### 3.2 Real Data Results

Figure 7 shows two Manhattan plots of Z-scores for the association between 3939 IDPs and AD, controlling for 754 imaging confounds, using `PCA-OLS1` and `DESLA-MP`. For `DESLA-MP`, 22 IDP associations pass the Benjamini-Hochberg False Discovery Rate (FDR) multiple testing threshold (Benjamini and Hochberg, 1995), controlling the FDR at *α* = 0.05. 5 IDP associations pass the Bonferroni threshold for controlling the Family Wise Error Rate (FWER) at *α* = 0.05. For `PCA-OLS1`, the FDR and Bonferroni threshold are the same, and only one IDP association passes both thresholds. All 5 IDP associations that pass the Bonferroni threshold using `DESLA-MP` are negative associations between AD and hippocampal volumes. It is well known and established that the hippocampus is severely affected by AD, and that AD is associated with atrophy of the hippocampus (Halliday, 2017). This gives us further confidence in the suitability of `DESLA-MP` as a high-dimensional inference method in population health neuroimaging, as it has replicated a well known result that the baseline approach `PCA-OLS1`, fails to detect.

**Figure 7.**
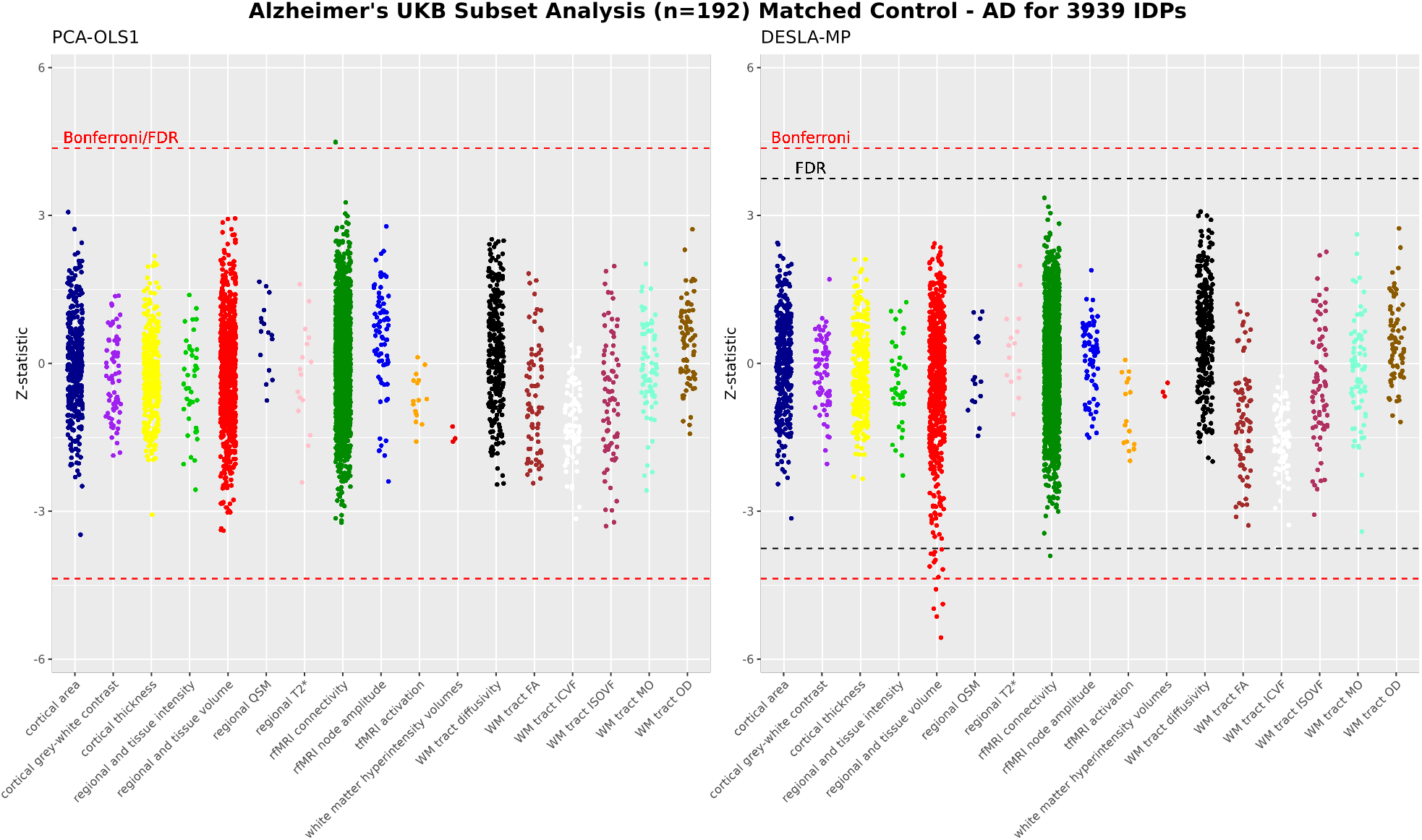
Z-statistic plots of associations between 3939 IDPs and AD, using `PCA-OLS1` (left) and `DESLA-MP` (right). `DESLA-MP` detects a number of negative associations between regional/tissue volumes and AD that `PCA-OLS1` fails to detect. The 5 IDPs that pass the Bonferroni threshold for `DESLA-MP` measure volumes in the hippocampus, which is severely affected by AD.

Figure 8 shows a Bland-Altman plot comparing the − log_10_ transformed p-values for all IDP on AD associations using `PCA-OLS1` and `DESLA-MP`. The plot does not show a clear, systematic tendency for `DESLA-MP` to have lower p-values than `PCA-OLS1`, but shows in the top right corner that there is a small number of associations for which `DESLA-MP` has significantly lower p-values. These are the regional and tissue volume IDPs that we found to have negative association with AD. This further raises our confidence in `DESLA-MP`, as it shows that the previously detected associations between IDPs and AD are unlikely to be the result of a systematic downward bias in p-values for `DESLA-MP`.

**Figure 8.**
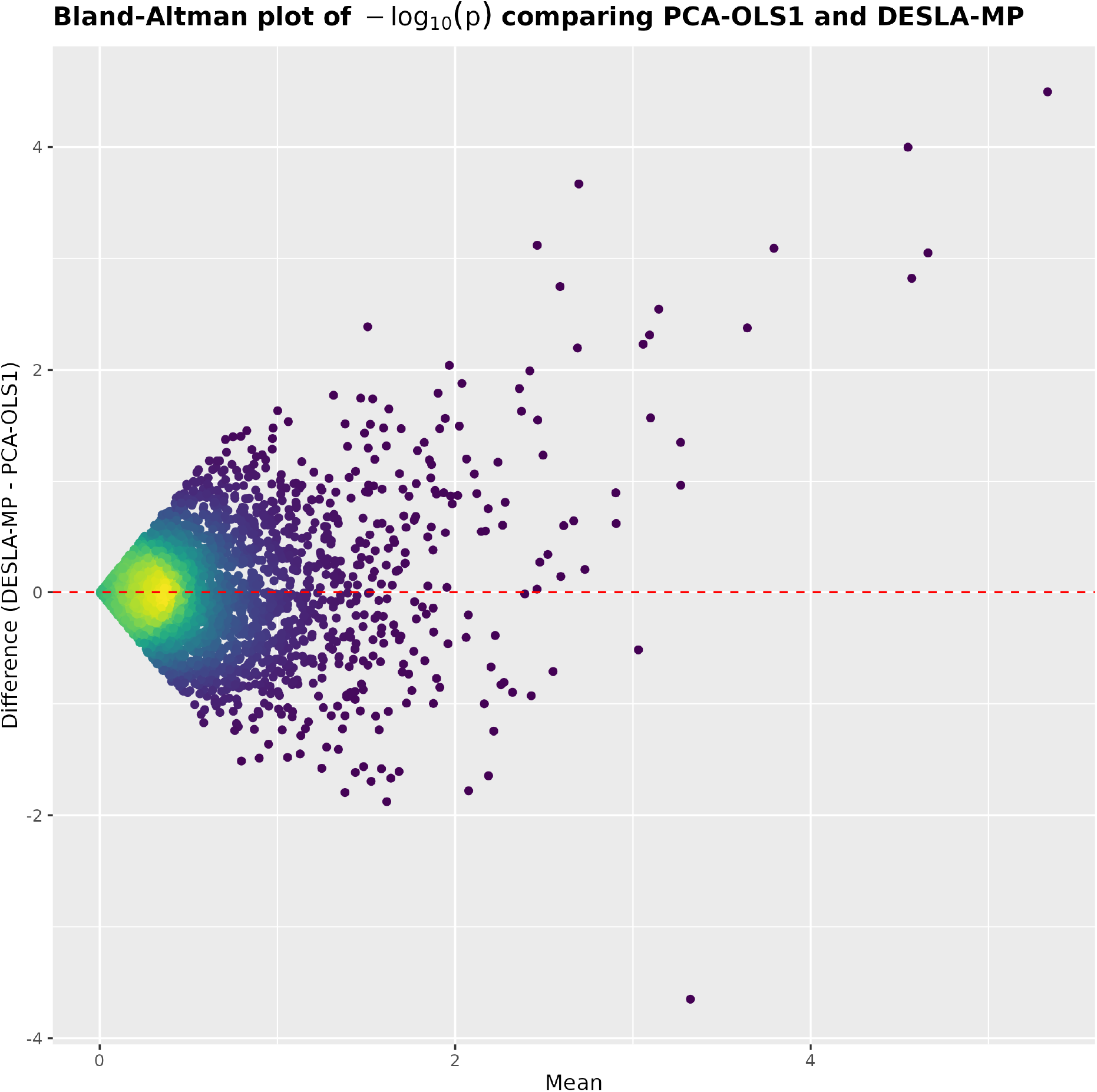
Bland-Altman plot comparing the − log_10_ transformed p-values of all IDP on AD associations using `PCA-OLS1` and `DESLA-MP`. We see no systematic bias in the p-values when comparing the two methods, but we see that there are a number of low p-values for `DESLA-MP` that are much higher for `PCA-OLS1`. These are the IDPs measuring hippocampal volumes, also seen in Figure 7.

Lastly, Figure 9 shows thw QQ-plots for the Z-scores of IDP on AD associations using `PCA-OLS1` and `DESLA-MP`. For `PCA-OLS1`, the QQ-plots show that the Z-scores are close to having a Gaussian distribution, which is indeed their true distribution under the null hypothesis of no association. For `DESLA-MP`, the highly negative Z-scores are more negative than expected under the null hypothesis. These Z-scores likely correspond to the IDPs that were found to have a negative association with AD. On the other hand, the highly positive Z-scores are less positive than expected under the null hypothesis, which is in agreement with our simulation results which show that `DESLA-MP` is conservative under the null hypothesis.

**Figure 9.**
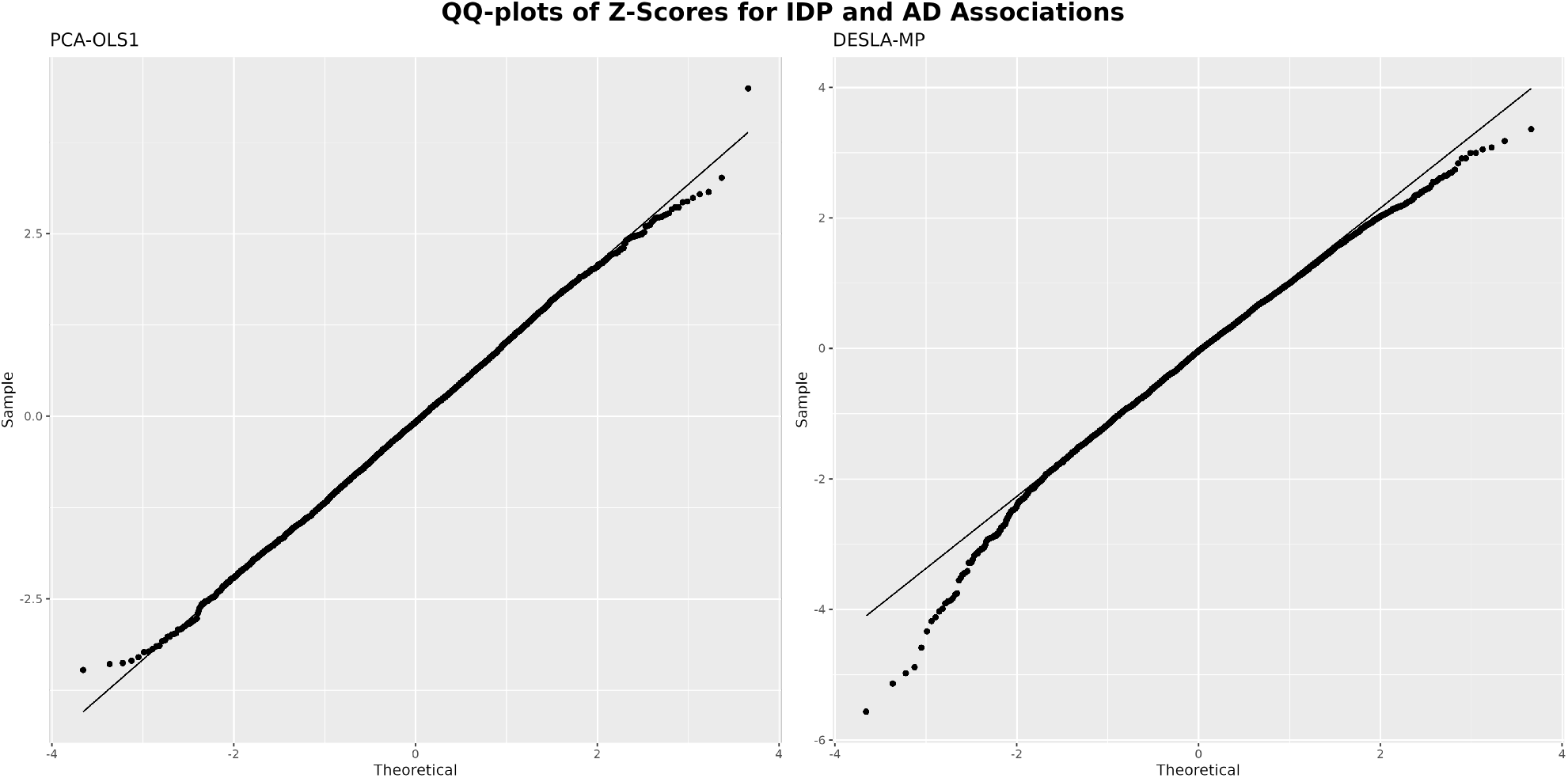
QQ-plots of the Z-scores corresponding to IDP on AD associations, for `PCA-OLS1` and `DESLA-MP`. We see that the Z-scores for `PCA-OLS1` look fairly Gaussian, which is their distribution under the null-hypothesis of no association. Conversely, for `DESLA-MP`, we see that the highly negative Z-scores are more negative than expected under the null-hypothesis, which corresponds to the hippocampal IDPs with strong negative associations with AD, and the highly positive Z-scores are less positive than expected under the null-hypothesis, which corresponds to the conservative FPR for `DESLA-MP` seen in the simulation study.

## 4 Discussion

We have evaluated 13 high-dimensional inference methods extensively in a simulation study, varying the sample size, strength of confounding and approximate sparsity of confounding effects. Unlike most previous studies on this topic, our simulation framework has assumed approximate sparsity, rather than strict sparsity, to give a more accurate picture of the performance on real data, which is not guaranteed to fall under exact sparsity. Overall, we have found that there is a number of methods that are reliable for the purposes of high-dimensional inference in UK Biobank population neuroimaging, even under conditions that are the most prone to induce false positives. Table 1 shows our recommendation for the method to use under different sample size scenarios, assuming that we are take a conservative approach of choosing the highest powered option among methods that we are highly confident have a controlled FPR. For example, we recommend `DESLA-MP` over `PCA-OLS2` for *n* = 500, even though both methods seem to have reasonably controlled FPRs at this sample size, and `PCA-OLS2` has higher power. This is because `PCA-OLS2` is shown to have an elevated FPR for higher sample sizes than *n* = 500, while `DESLA-MP` shows conservative FPRs overall.

**Table 1:** The recommended methods for high-dimensional inference in the UK Biobank Brain Imaging Cohort at different sample sizes. At the lowest sample size, *n* = 50, we recommend a ridge regression permutation test based on the Freedman-Lane proceedure. For sample sizes ranging between *n* = 100 and *n* = 500, we recommend a de-sparsified LASSO approach using the Moore-Penrose inverse for approximating the precision matrix of our data. For *n* ≥ 1000, we recommend `PCA-OLS1`, which is equivalent to vanilla OLS at this sample size. The recommendations are based on the results of our simulation study.

| Sample Size | Recommended Method |
| --- | --- |
| $n = 50$ | Ridge.PT.FLH1 |
| $n = 100$ | DESLA-MP |
| $n = 200$ | DESLA-MP |
| $n = 500$ | DESLA-MP |
| $n = 1000$ | PCA-OLS1 |
| $n = 2000$ | PCA-OLS1 |

The main limitation of our work is that we have only evaluated the FPR and power of our methods using confidence level *α* = 0.05, but the confidence level *α* = 0.05 is a somewhat arbitrary threshold, and much lower thresholds are often required, especially when correcting for multiple testing. A simulation study evaluating the FPR and power of all methods for some confidence level *α* << 0.05 will necessarily require a substantially larger number of simulations, and will due to computational constraints therefore necessitate a simulation framework that is more focussed on a few specific methods and settings.

We have created a user-friendly `R` package, `HDConf`, which includes all of the methods included in this work. The package is freely available for download on GitHub: https://github.com/lavrad99/HDConf

## Data Availability

Not applicable

## Declarations

### Ethics approval and consent to participate

The UK Biobank has received ethical approval from the North West Multi-Center Research Ethics Committee (11/NW/0382).

This research project received approval from the UKB under application number 8107.

This research project has adhered to the Declaration of Helsinki.

### Consent for publication

Not applicable.

### Availability of data and materials

All code can be found in the following repository: https://github.com/lavrad99/HDConf

## Acknowledgements

The computational aspects of this research were supported by the Wellcome Trust Core Award Grant Number 203141/Z/16/Z and the NIHR Oxford BRC. The views expressed are those of the author(s) and not necessarily those of the NHS, the NIHR or the Department of Health

## 5 Supplementary Material

**Figure S1:**
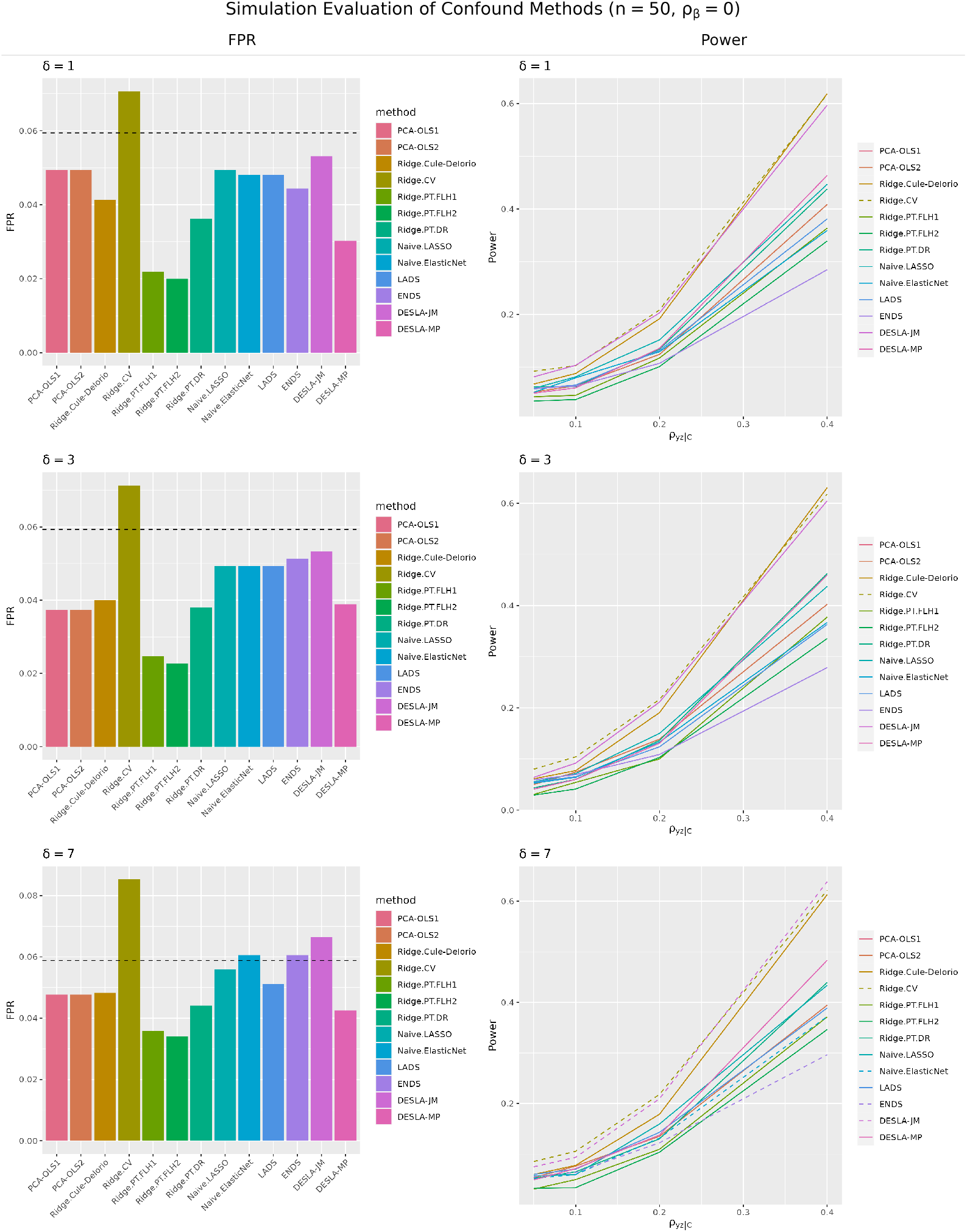
Simulation results for *n* = 50, *ρ*_*β*_ = 0. The bar plots to the left show the FPR and lineplots the statistical power of different methods. The dashed line in the bar plots is the 95% upper confidence limit for a FPR of 0.05. Methods above this limit have dashed lines in the power plot.

**Figure S2:**
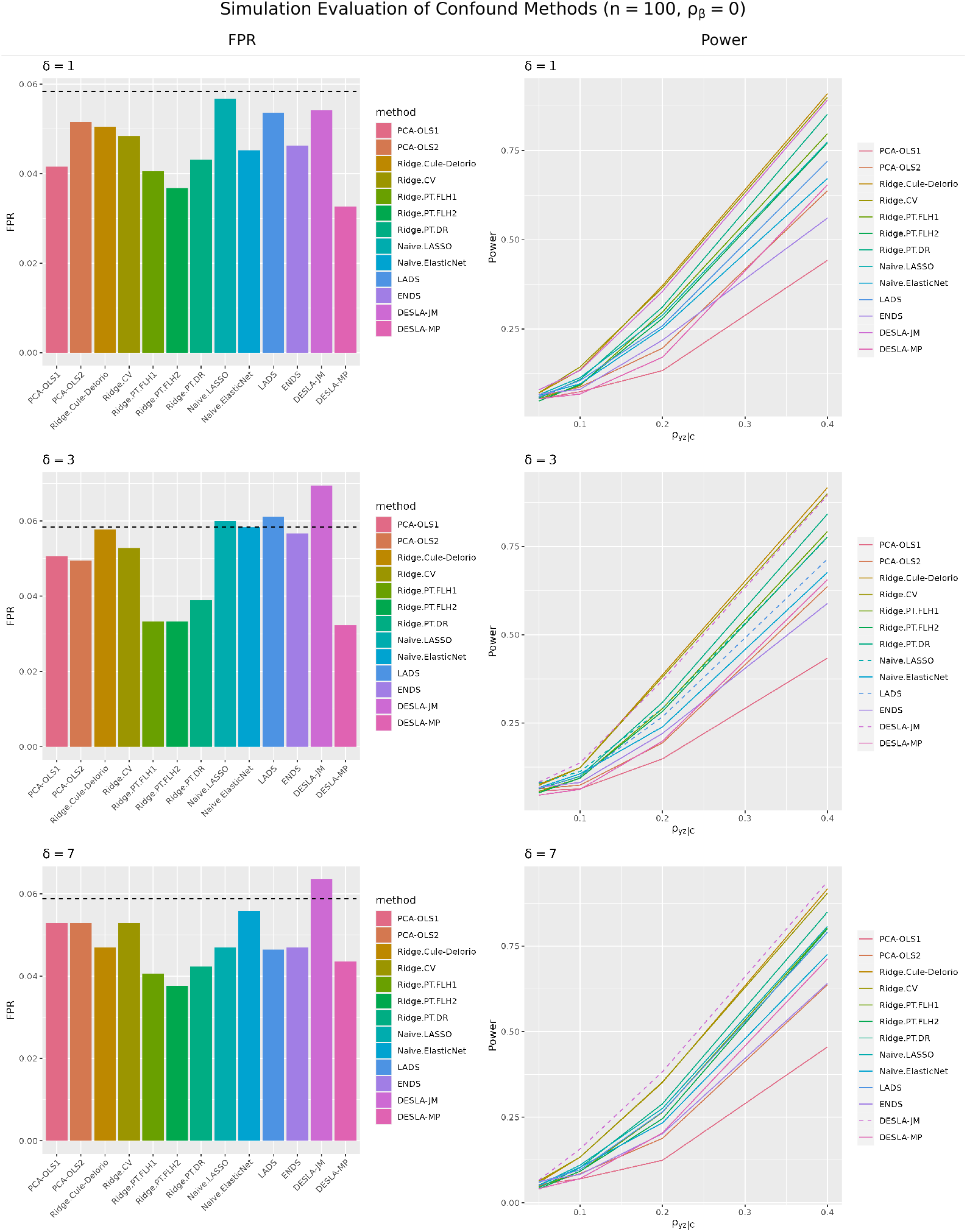
Simulation results for *n* = 100, *ρ*_*β*_ = 0. The bar plots to the left show the FPR and lineplots the statistical power of different methods. The dashed line in the bar plots is the 95% upper confidence limit for a FPR of 0.05. Methods above this limit have dashed lines in the power plot.

**Figure S3:**
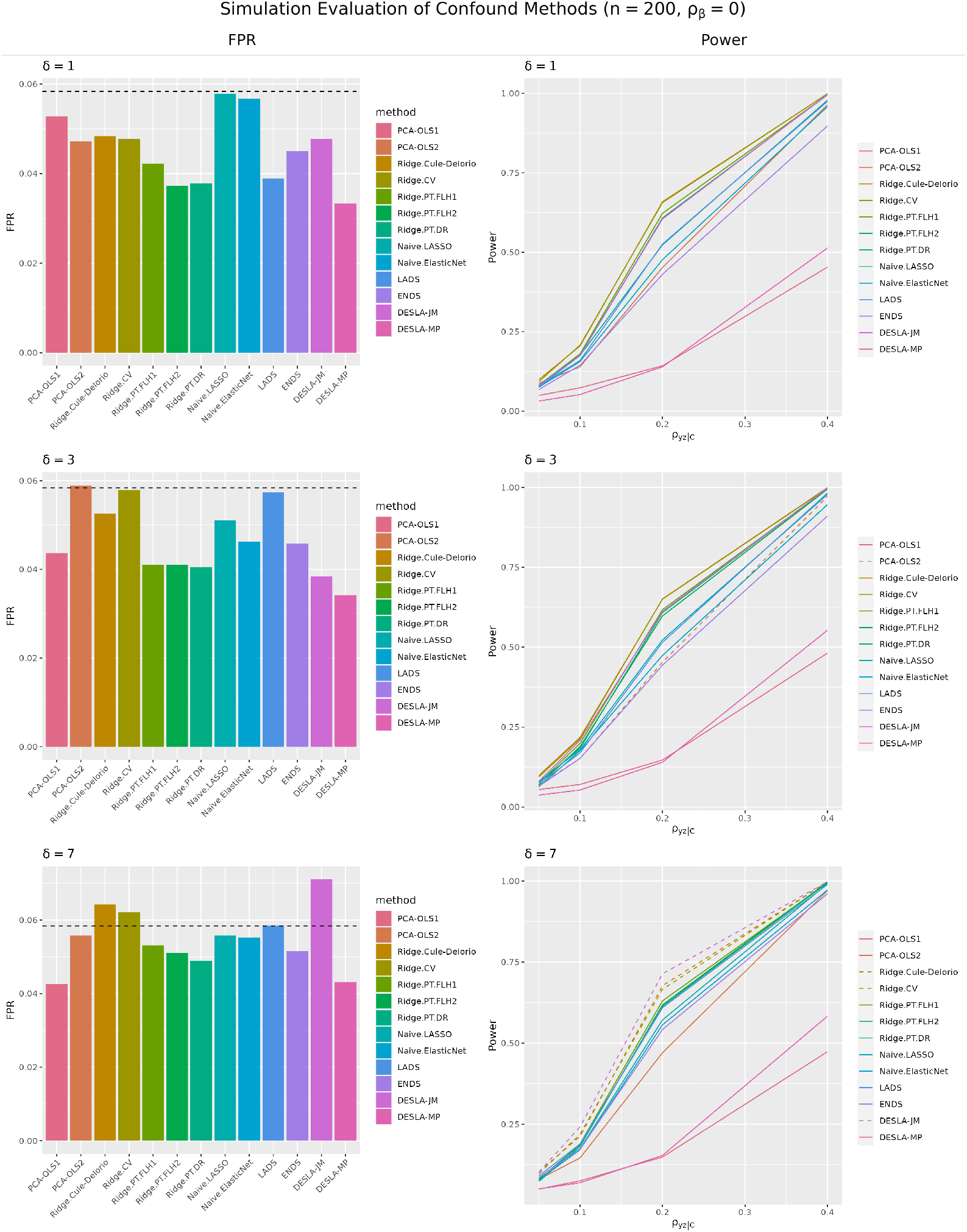
Simulation results for *n* = 200, *ρ*_*β*_ = 0. The bar plots to the left show the FPR and lineplots the statistical power of different methods. The dashed line in the bar plots is the 95% upper confidence limit for a FPR of 0.05. Methods above this limit have dashed lines in the power plot.

**Figure S4:**
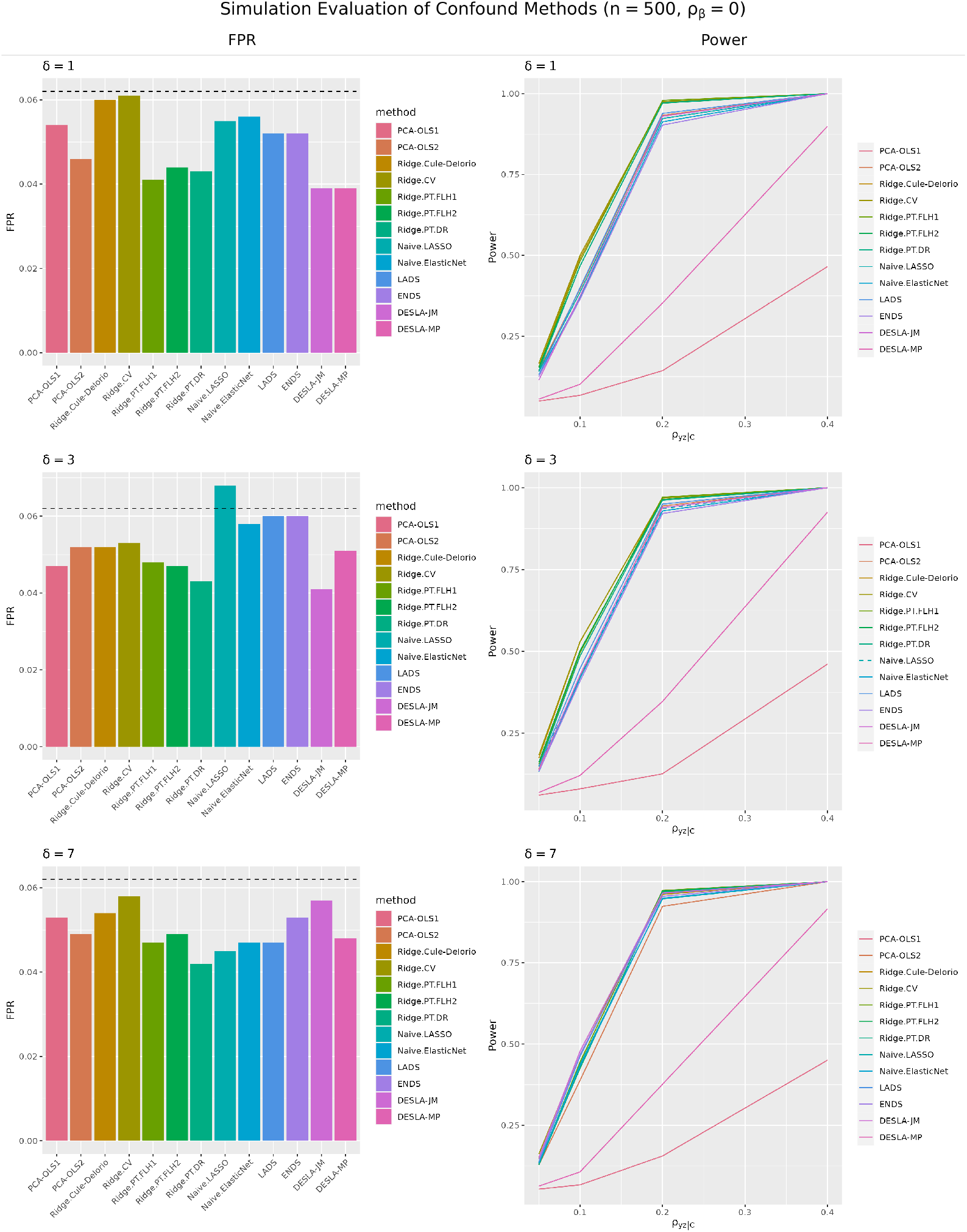
Simulation results for *n* = 500, *ρ*_*β*_ = 0. The bar plots to the left show the FPR and lineplots the statistical power of different methods. The dashed line in the bar plots is the 95% upper confidence limit for a FPR of 0.05. Methods above this limit have dashed lines in the power plot.

**Figure S5:**
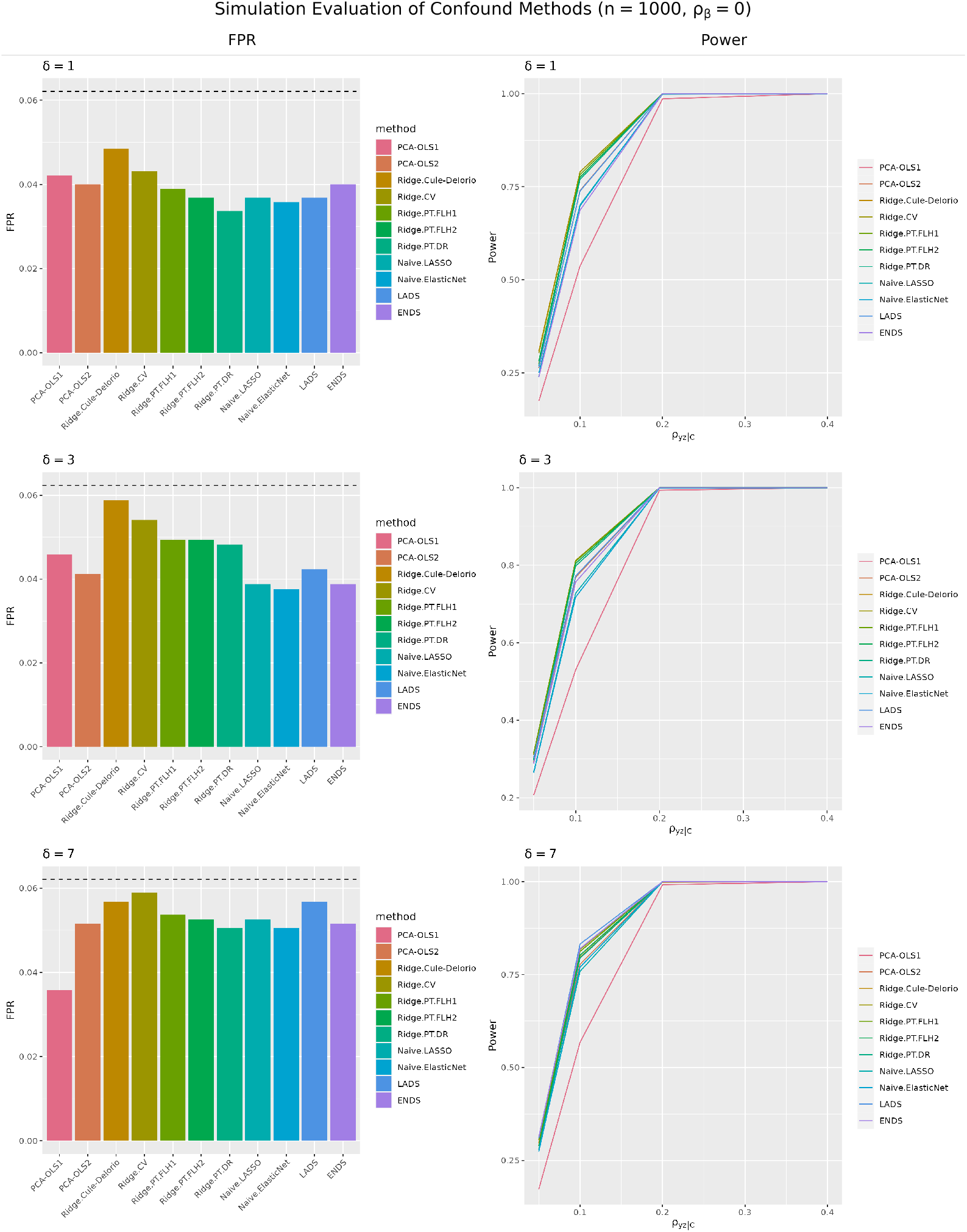
Simulation results for *n* = 1000, *ρ*_*β*_ = 0. The bar plots to the left show the FPR and lineplots the statistical power of different methods. The dashed line in the bar plots is the 95% upper confidence limit for a FPR of 0.05. Methods above this limit have dashed lines in the power plot.

**Figure S6:**
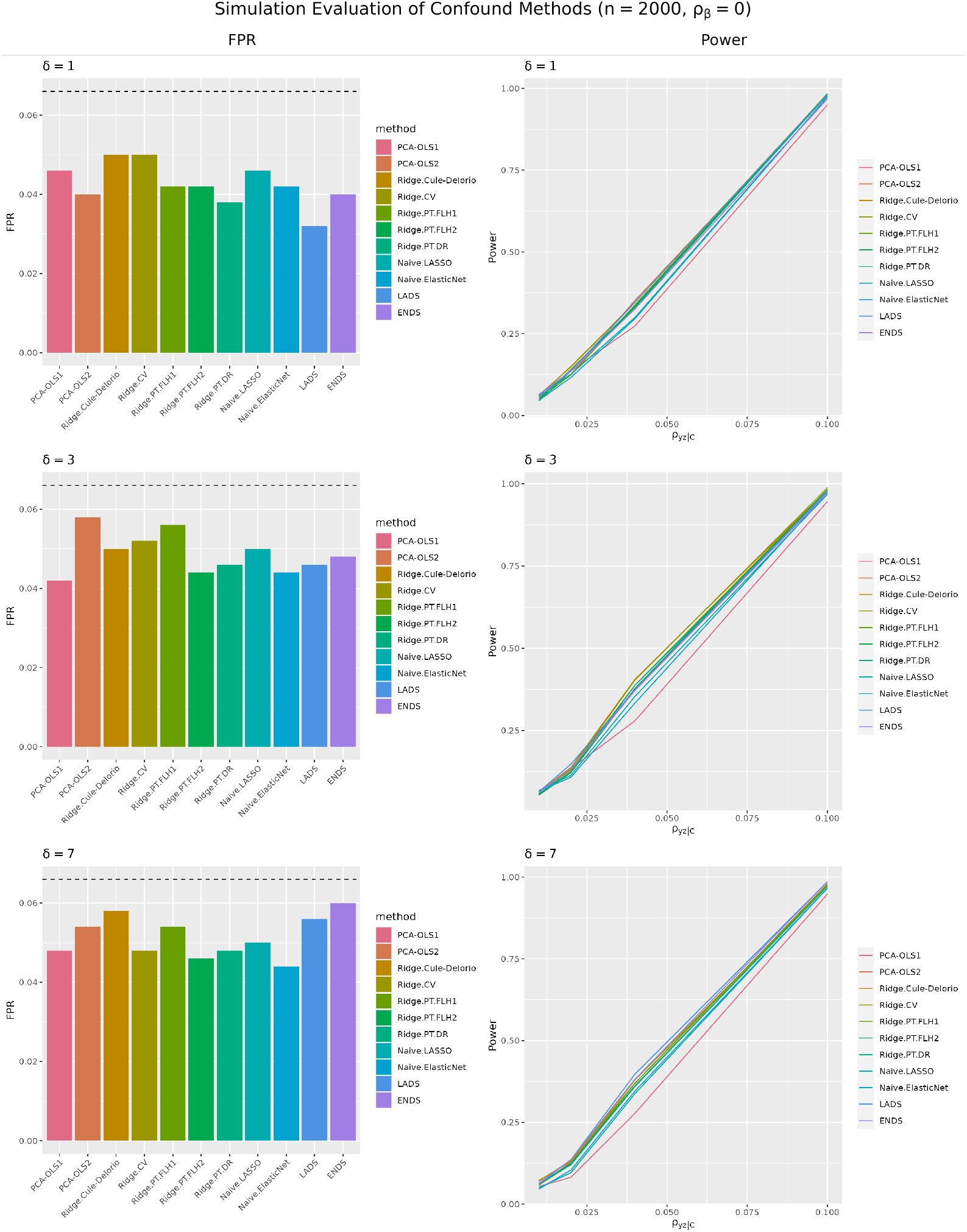
Simulation results for *n* = 2000, *ρ*_*β*_ = 0. The bar plots to the left show the FPR and lineplots the statistical power of different methods. The dashed line in the bar plots is the 95% upper confidence limit for a FPR of 0.05. Methods above this limit have dashed lines in the power plot.

**Figure S7:**
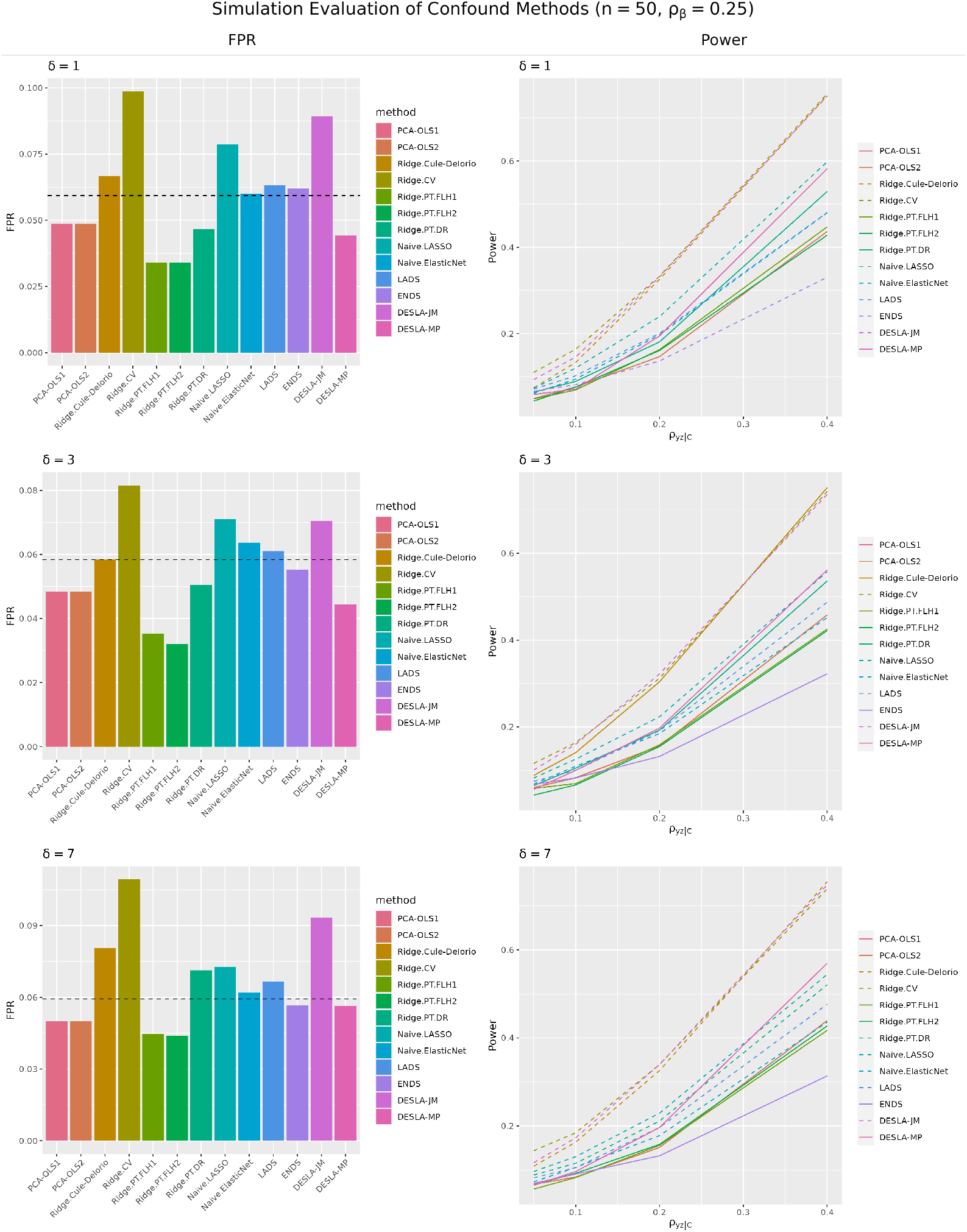
Simulation results for *n* = 50, *ρ*_*β*_ = 0.25. The bar plots to the left show the FPR and lineplots the statistical power of different methods. The dashed line in the bar plots is the 95% upper confidence limit for a FPR of 0.05. Methods above this limit have dashed lines in the power plot.

**Figure S8:**
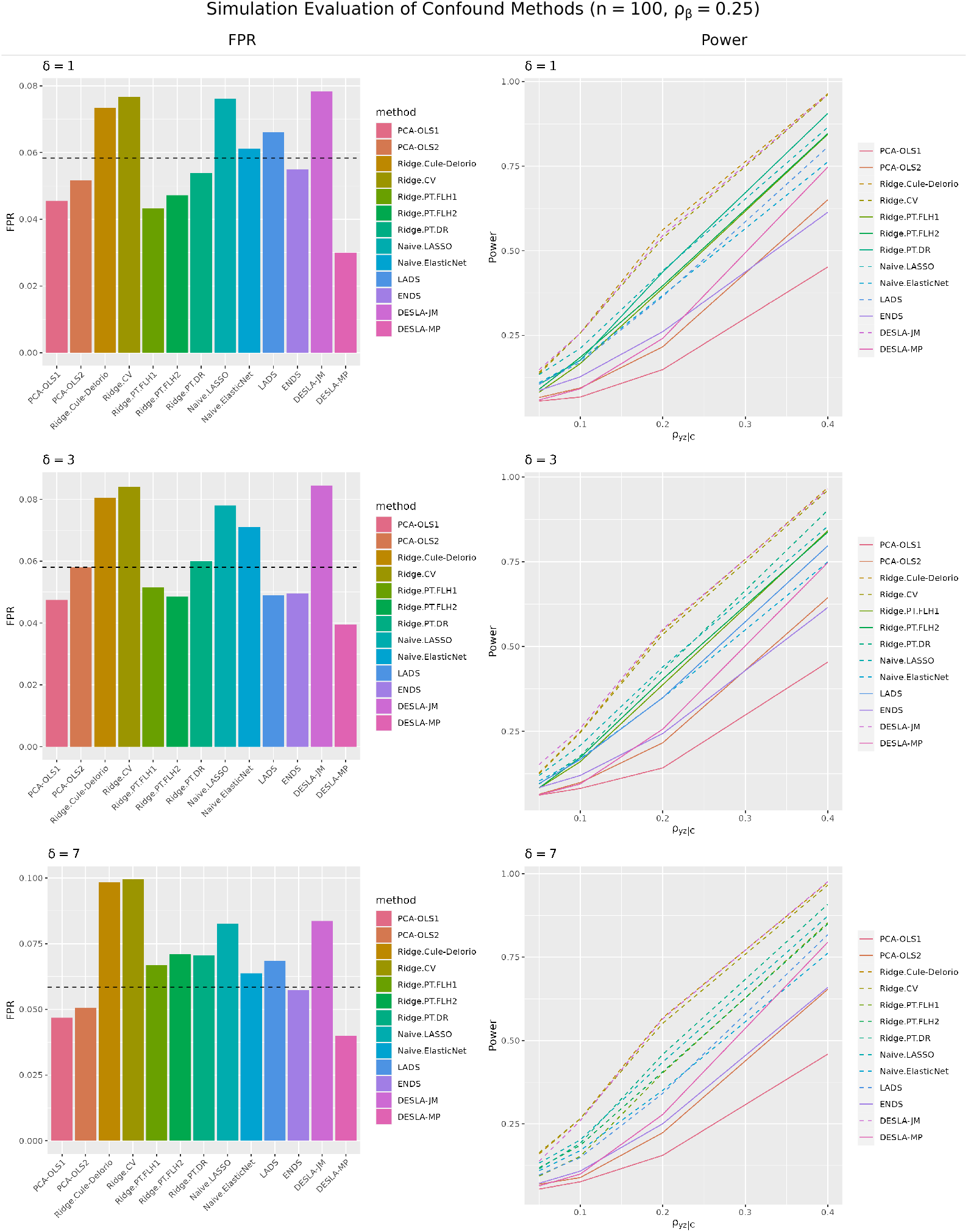
Simulation results for *n* = 100, *ρ*_*β*_ = 0.25. The bar plots to the left show the FPR and lineplots the statistical power of different methods. The dashed line in the bar plots is the 95% upper confidence limit for a FPR of 0.05. Methods above this limit have dashed lines in the power plot.

**Figure S9:**
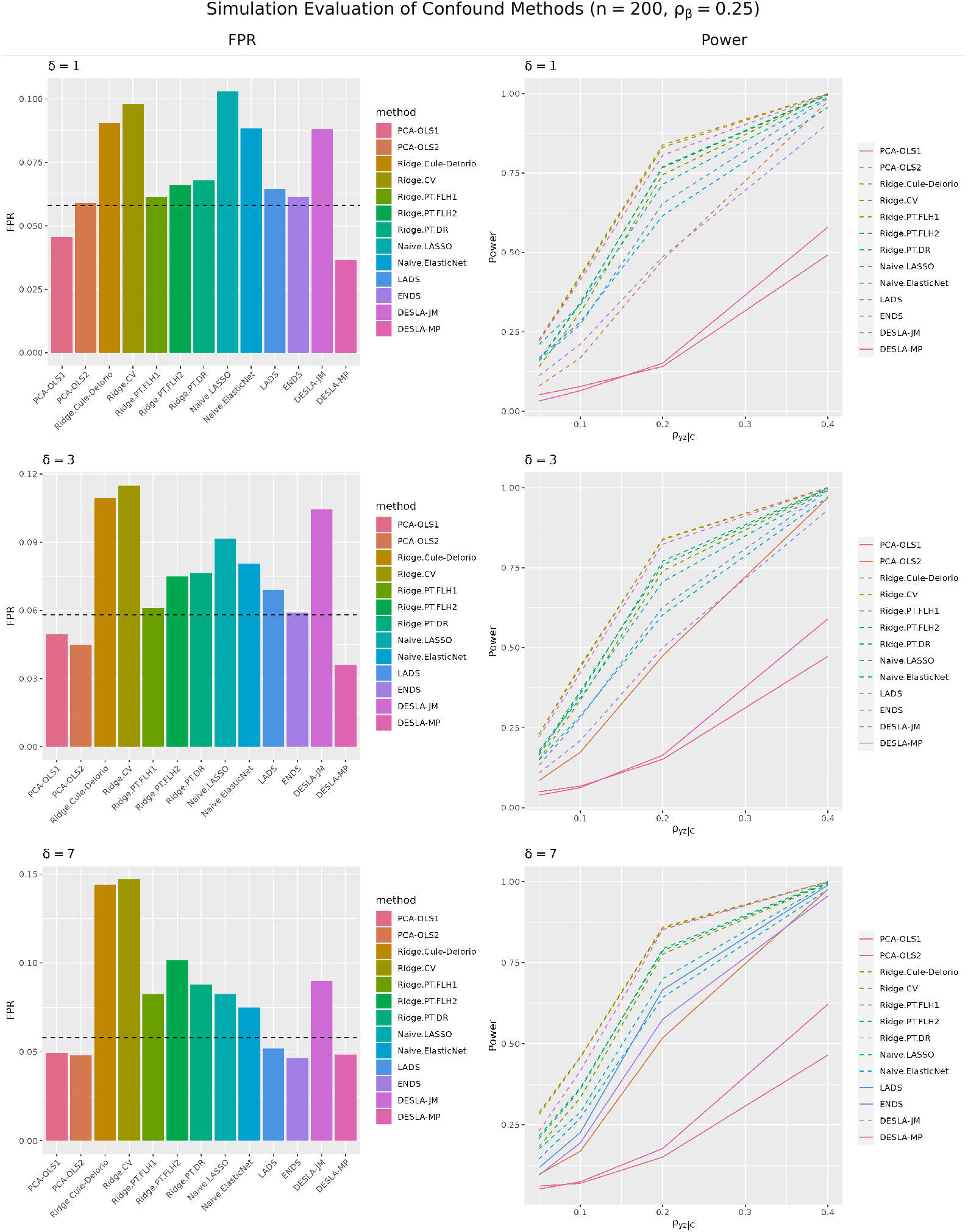
Simulation results for *n* = 200, *ρ*_*β*_ = 0.25. The bar plots to the left show the FPR and lineplots the statistical power of different methods. The dashed line in the bar plots is the 95% upper confidence limit for a FPR of 0.05. Methods above this limit have dashed lines in the power plot.

**Figure S10:**
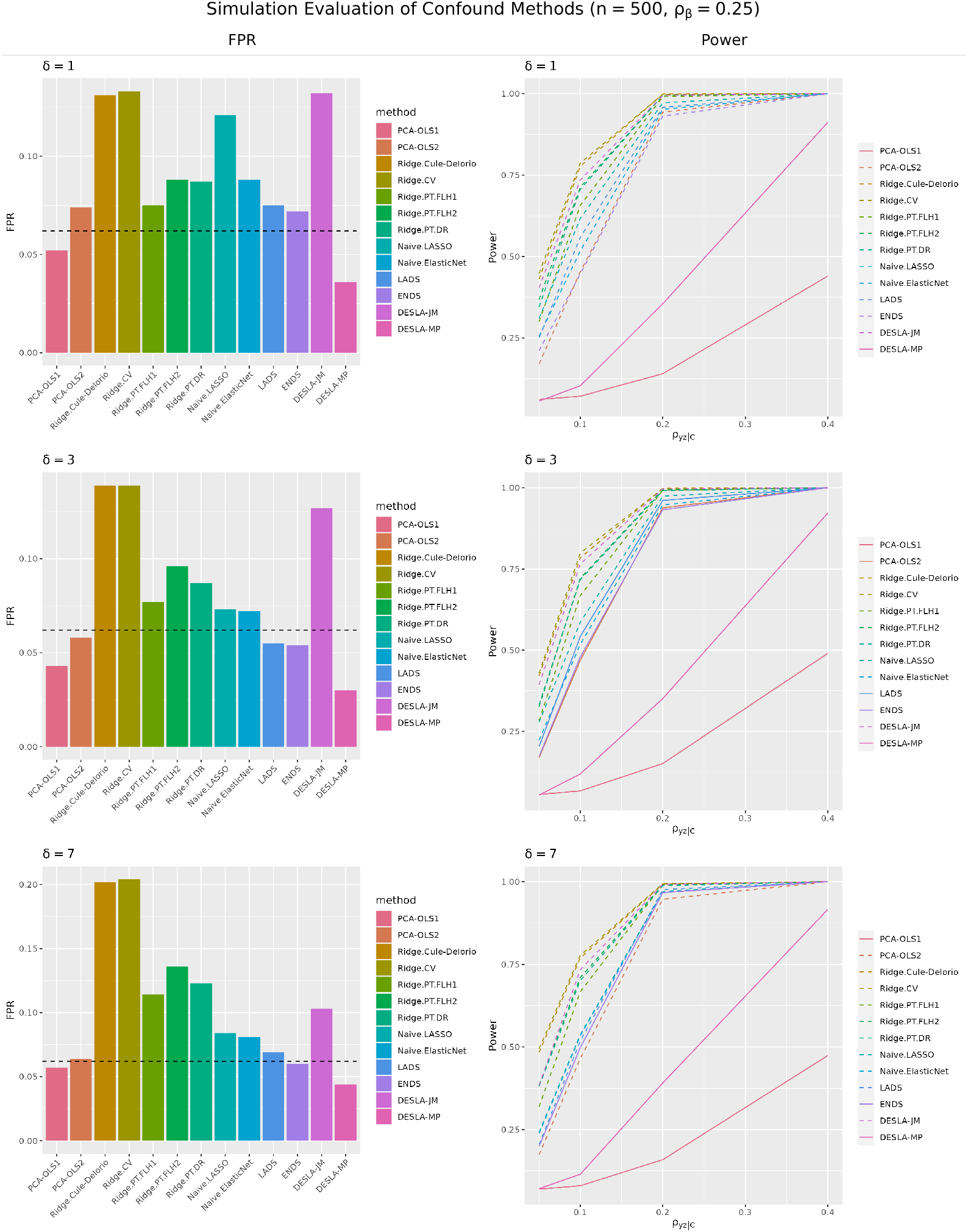
Simulation results for *n* = 500, *ρ*_*β*_ = 0.25. The bar plots to the left show the FPR and lineplots the statistical power of different methods. The dashed line in the bar plots is the 95% upper confidence limit for a FPR of 0.05. Methods above this limit have dashed lines in the power plot.

**Figure S11:**
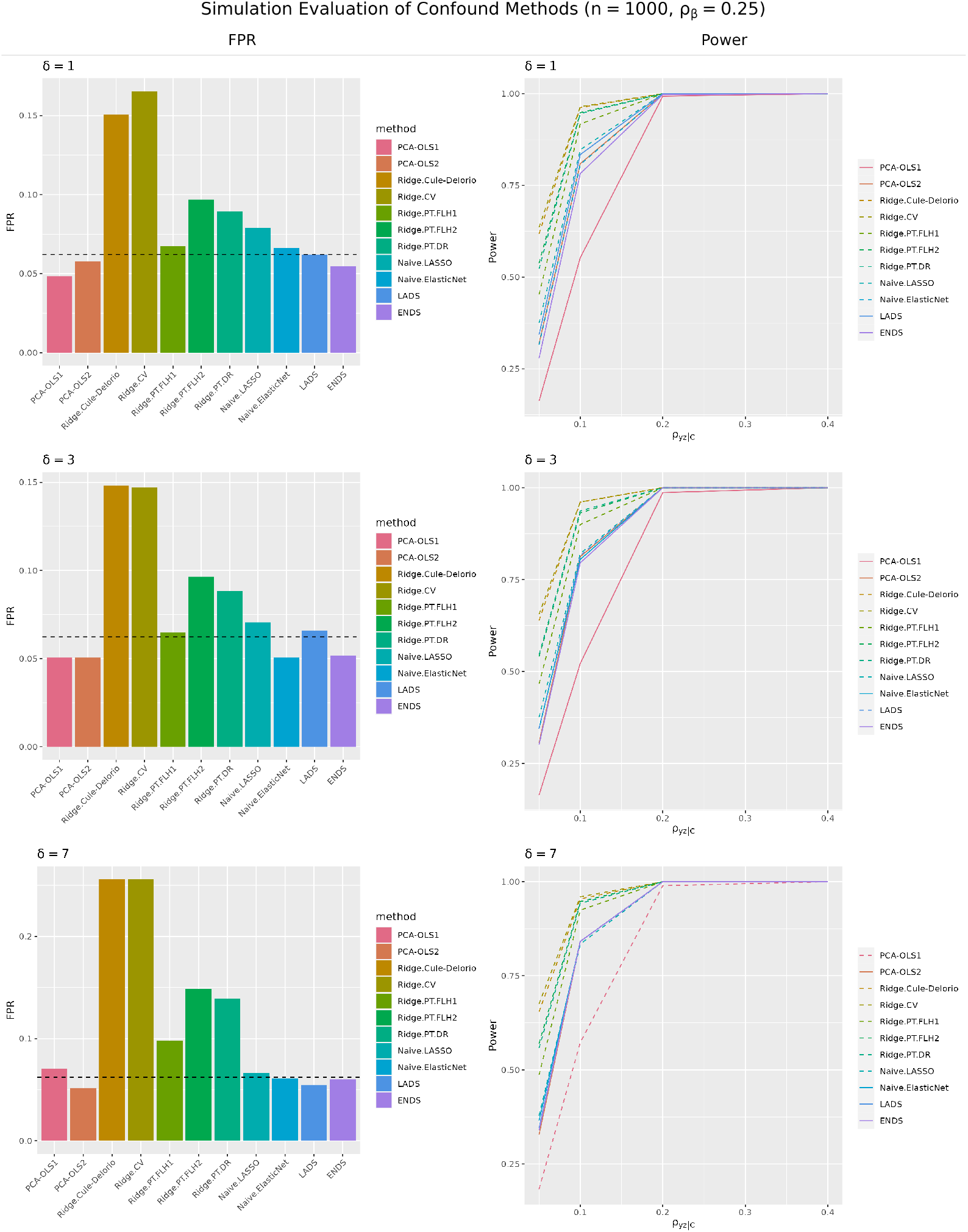
Simulation results for *n* = 1000, *ρ*_*β*_ = 0.25. The bar plots to the left show the FPR and lineplots the statistical power of different methods. The dashed line in the bar plots is the 95% upper confidence limit for a FPR of 0.05. Methods above this limit have dashed lines in the power plot.

**Figure S12:**
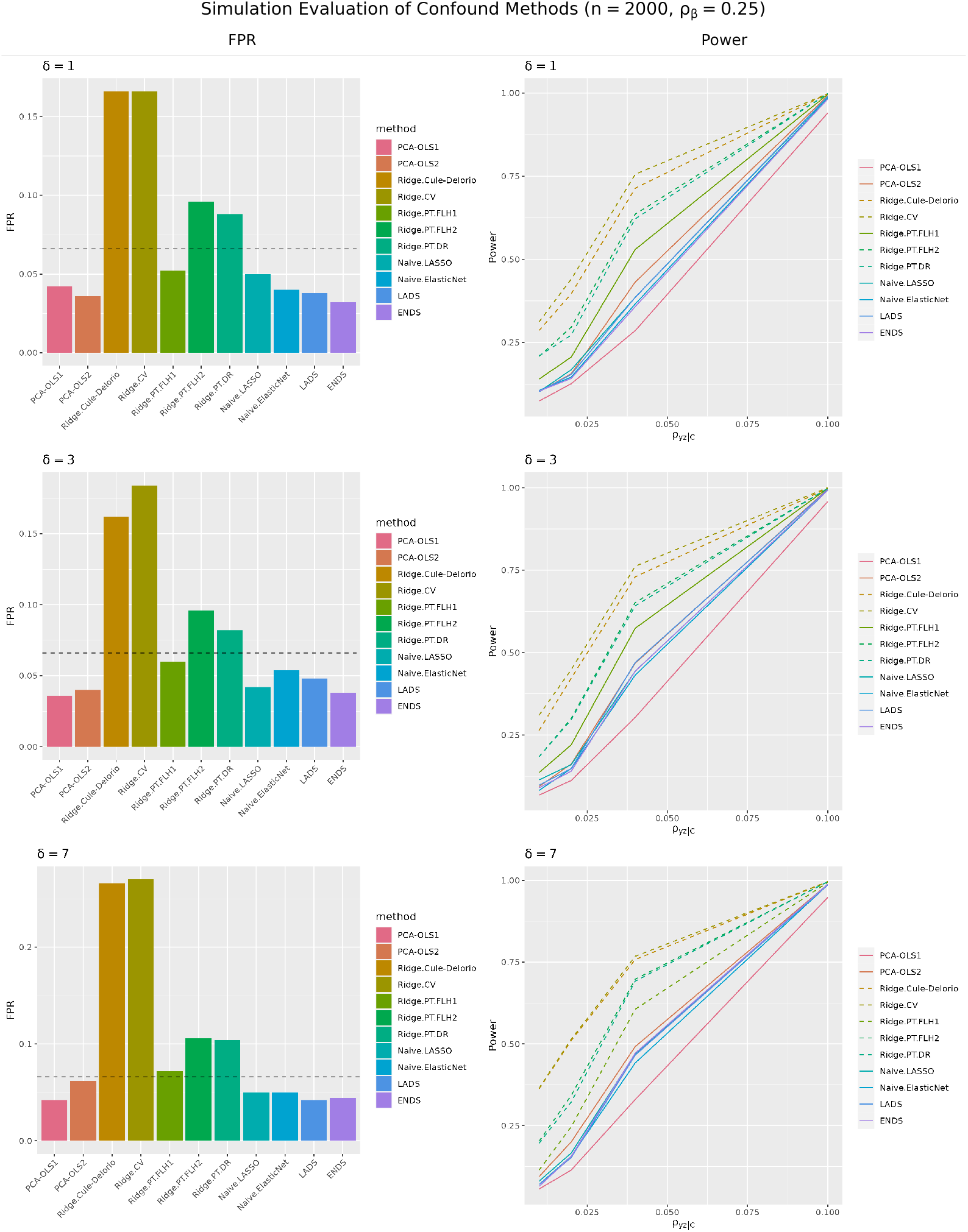
Simulation results for *n* = 2000, *ρ*_*β*_ = 0.25. The bar plots to the left show the FPR and lineplots the statistical power of different methods. The dashed line in the bar plots is the 95% upper confidence limit for a FPR of 0.05. Methods above this limit have dashed lines in the power plot.

